# Predictive performance of wearable sensors for mortality risk in older adults: a model development and validation study

**DOI:** 10.1101/2025.04.03.25325101

**Authors:** Charlie Harper, Adam D. Sturge, Shing Chan, Ben Maylor, Alaina Shreves, Daniel Meier, Prachi Patkee, John Schoonbee, Adam Strange, Christoph Nabholz, Derrick Bennett, Aiden Doherty

## Abstract

**Background:** Many adults in high-income countries carry a device capable of measuring physical-activity behaviour. Thus, there is public health need to understand whether such data can enhance prediction of future health outcomes. We aimed to investigate whether device-measured daily-step count and walking cadence improve the prediction of mortality beyond traditional risk-factors.

**Methods:** Risk models were developed to predict five-year all-cause mortality using data from the UK Biobank accelerometer sub-study, with external validation in the US 2011-2014 National Health and Nutrition Examination Survey (NHANES). Median daily-step count and peak one-minute walking cadence were derived using self-supervised machine learning models from seven-day wrist-worn accelerometer data. Cox models were used to develop a baseline model incorporating traditional risk-factors, and a baseline model plus accelerometer data (i.e. daily-steps and walking cadence). Changes in model performance were assessed using Harrell’s C-index, net reclassification index (NRI; 10% threshold), and the Nam-D’Agostino calibration test.

**Findings:** Among 79,717 UK Biobank participants, 1,640 died within 5-years. Adding accelerometer data to the baseline model modestly improved risk discrimination and classification with a change in c-index of 0.008 (95% confidence interval [CI] 0.005-0.011) and 3.3% NRI (95%CI 2.1%-4.5%). Greatest improvements in prediction were observed in participants with prior disease at baseline, showing a change in c-index of 0.028 (95%CI 0.019-0.039) and 5.9% NRI (95%CI 3.1%-8.6%). In the NHANES external validation cohort (n=4,713; deaths=378), similar improvements in prediction were observed (change in c-index: 0.015, 95%CI 0.007-0.025; NRI: 4.0%, 95%CI 0.7%-7.4%). All models were well calibrated (Nam-D’Agostino χ^2^ range: 6.8-13.2).

**Interpretation:** Device-measured daily-step count and walking cadence consistently demonstrated modest improvements in predicting mortality risk beyond traditional risk-factors, with the most significant enhancements seen in individuals with prior disease. These findings suggest that incorporating information from wearables does provide important new ways to improve risk stratification for targeted intervention in high-risk individuals.

## Introduction

Risk prediction models are an essential public health tool that enable the targeting of interventions at those most at risk of morbidity and mortality.^1^ Such prediction models typically rely on information about traditional risk factors collected during an individual’s interaction with their healthcare system, including: age, sex, self-reported cigarette and alcohol use, disease history, physiological and biological measurements.^2,3^ Yet, perhaps due to technological limitations, these models have overlooked habitual behaviours occurring outside of the clinical setting (i.e. in a free-living environment) that may play an important role in determining a person’s risk. In the past decade there has been growing evidence of a strong dose-response relationship between daily levels of physical activity and mortality.^4,5^ With most adults in high-income countries now carrying digital devices that can continuously measure physical activity behaviours, there is a public health need to understand whether such data can provide important improvements in predicting future health outcomes.^6,7^

Several studies have explored the predictive performance of device-measured physical activity behaviours, finding modest improvements in mortality prediction when added to models with traditional risk factors.^8–13^ For instance, Chen et al^8^ found that incorporating over 20 aspects of an individual’s physical activity, such as overall activity, duration, frequency, and intensity distribution, led to important improvements in prediction. However, these studies often had limited sample sizes and focused on measures of physical activity behaviour not reported by consumer-grade devices or understood by the general public^14^, thus limiting their application in current public health decision making.

To address these limitations, we aimed to investigate two physical activity metrics routinely reported by consumer-grade devices and easily understood by the public: daily step count and walking cadence., defined as the number of steps taken per minute. Using data from two prospective cohorts, we developed and validated a novel risk prediction algorithm for 5-year all-cause mortality. We aimed to determine whether device-measured daily step count and walking cadence improve prediction of 5-year mortality risk over and above traditional risk factors, including demographic and behavioural variables, prior disease, and common laboratory measures (Supplemental table 2-3). A secondary objective was to identify which sub-populations yielded the greatest improvements in prediction, allowing for a more targeted use of wearable-device data in risk models.

## Methods

### Model development cohort

The UK Biobank is a population-based prospective cohort study of over 500,000 participants in England, Scotland and Wales, recruited between 2006 and 2010.^15^ At baseline, participants attended an assessment involving a touchscreen questionnaire, biological sampling, an interview by a trained interviewer, and anthropometric measurements. Between June 2013 and December 2015, over 230,000 participants with a valid email address were invited to wear a accelerometer.^16^ Over 100,000 consenting participants were mailed an Axivity AX3 wrist-worn triaxle accelerometer at 100Hz to be worn on the dominant wrist for 7 days. The median time between baseline assessment and accelerometer wear was 5.7 years (interquartile range [IQR] 4.9-6.5). The UK Biobank has ongoing passive follow-up via linkage to routinely collected data sources, including death registry data obtained from NHS England for participants in England and Wales, and NHS Scotland Central Register (up to November 30 2022). These linked data include information on date of death, underlying (i.e. primary cause) and other contributing causes of death. Participants in the UK Biobank provided written informed consent. The study was approved by the National Information Governance Board for Health and Social Care and the National Health Service North West Multicentre Research Ethics Committee (06/MRE08/65).

### External validation cohort

The National Health and Nutrition Examination Survey (NHANES) is a large, ongoing, cross-sectional study of the non-institutionalised United States population collected annually and made publicly available by the Centers for Disease Control.^17^ Consenting participants undergo an interview, biological sampling, and anthropometric measurements. During the 2011-14 rounds of data collection, participants aged ≥3 years were invited to wear an ActiGraph GT3X+ wrist-worn triaxle accelerometer at 80Hz to be worn on the non-dominant wrist over 9 calendar days (of which the first and last days were partial days), with up to 7 full days of data.^18^ Baseline assessment data were collected at a mobile examination centre immediately prior to accelerometer wear, with participants asked to return the device via mail at the end of the study. NHANES has ongoing passive follow-up for participants via linkage to the National Center for Health Statistics public-use linked mortality files (up to December 31 2019). These linked data include information on date of death and underlying cause of death.

### Measurements derived from wrist-worn accelerometer data

A previously validated open-source self-supervised machine learning algorithm (OxWearables “stepcount” package version 3.9.0; https://github.com/OxWearables/stepcount) was applied to each participant’s raw accelerometer data in the UK Biobank and NHANES, extracting the median total daily step count and the median daily peak one-minute bout of steps (i.e. walking cadence).^19^ These two metrics together are termed “accelerometer data”. Missing periods due to non-wear, defined by acceleration standard deviations <13mg, were imputed by averaging the step counts in the corresponding times across valid days.^16,20^ To ensure adequate accelerometer data quality: (1) the first and last days of accelerometer wear and days with less than 16 hours of wear were excluded for participants in the NHANES; and (2) participants in either cohort were excluded if the algorithm failed, if there was poor calibration, insufficient wear time (<3 days), unrealistic acceleration (>100mg), or overall step values equal to zero.

### Outcome

The primary outcome was all-cause mortality at five years, ascertained through record linkage. The secondary outcome was cause-specific mortality, where the underlying cause of death (coded using the International Classification of Diseases, 10^th^ Revision [ICD-10]) was categorised as vascular (UK Biobank: I00-I199; NHANES: I00-I09, I11, I13, I20-I51 and I60-I69), cancer (C00-C97), or other causes of death (i.e., not attributed to vascular/cancer).

### Statistical analyses

Participants were followed-up starting the day after accelerometer wear to the date of death or censoring at five years, whichever came first. Participants missing covariate data were excluded in a complete case analysis. For NHANES, participants were also excluded if they were younger than 40 years at the time of accelerometer wear or had less than five years of follow-up. Sample size calculations estimated that there were enough events in the UK Biobank and the NHANES to develop and validate prediction models with up to 40 covariates (**Supplemental Table 1**).^21^

Using data from the UK Biobank, four models were developed: a baseline model, baseline model plus accelerometer data, laboratory model, and laboratory model plus accelerometer data.

The baseline model covariates included established traditional risk factors used in previous studies investigating the predictive performance of accelerometer data for mortality.^8–13^ The covariates included 5-year age group at accelerometer wear (<50, 50-54, 55-59, 60-64, 65-69, 70-74, and ≥75 years), sex (male or female), current smoking status (yes or no), body mass index (BMI; kg/m^2^), systolic blood pressure (mmHg), whether participants were taking medication to manage blood pressure or cholesterol (yes or no), and prior disease (including myocardial infarction, stroke, congestive heart failure, cancer and diabetes; **Supplemental Tables 2-4**).

The laboratory model included risk factors from the baseline model plus a range of laboratory measures found to be most predictive of mortality in the UK Biobank using two criteria: (1) significantly associated with mortality (p-value <0.05) in a multivariable Cox model, and (2) not highly correlated with other measures (Pearson’s correlation coefficient <0.7). The additional laboratory covariates included: albumin (g/L), alanine aminotransferase (U/L), aspartate aminotransferase (U/L), high-density lipoprotein cholesterol (mmol/L), gamma-glutamyltransferase (U/L), lymphocyte count (10^9 cells/L), mean corpuscular haemoglobin (pg), mean corpuscular haemoglobin concentration (g/dL), red blood cell count (10^12 cells/L), red blood cell distribution width (%), urea (mmol/L), and white blood cell count (10^9 cells/L). Low-density lipoprotein (LDL) cholesterol was not included in the variable selection process due to a high degree of missingness in NHANES, were only a small subset of participants were selected for LDL measurement (36.5%).

The baseline model plus accelerometer data and laboratory model plus accelerometer data included the covariates set out above, as well as median daily step count and walking cadence. Cox proportional hazards models were used to estimate the coefficients for the four prediction models, and fractional polynomial terms (selected using a multivariable model) were used to model continuous covariates exhibiting non-linear relationships with mortality risk.^22^ All risk factors were mean centred prior to model fitting.

These four models were then externally validated in the NHANES cohort, where model coefficients and baseline hazard were updated. This was due to two key disparities between the cohorts: (1) prior disease information was ascertained by self-report only in NHANES, thus the opportunity for false positives was much higher; and (2) due to the greater proportion of participants ≥75 years in the NAHNES cohort the five-year mortality rates were almost four times higher than in the UK Biobank.

Discriminative performance was assessed using Harrell’s c-index with 95% confidence intervals [CI] generated using 1,000 bootstrap samples and corrected for optimism.^23^ The additive value of accelerometer data was assessed by calculating the change in c-index between the baseline and laboratory models versus the same model plus accelerometer data. Reclassification performance was evaluated using integrated discrimination improvement (IDI), and net reclassification index (NRI) at the 10% risk treshold.^24,25^ Calibration plots were generated, presenting deciles of five-year predicted risk with smoothed calibration curves to show overall calibration. Calibration performance was measured using the Nam-D’Agostino test (with 9 degrees of freedom) to derive a chi-squared statistic for each model.^26^

Sensitivity analyses investigated whether there was heterogeneity in discriminative and recalibration performance by sex (male, female), age group (<70 years, ≥70), prior disease (yes, no), and cause of death (vascular, cancer, or other). Analyses were conducted using R version 4.2.1 and RStudio version 2022.07. This study complies with TRIPOD guidance on the reporting of risk prediction studies (see **Supplemental Table 5** for checklist).^27^

## Results

### Baseline characteristics

In the UK Biobank and NHANES cohorts, 79,717 and 4,713 participants were included in the analyses, respectively (**Supplemental Figures 1 and 2**). Median age at time of accelerometer wear was 63 years (interquartile range [IQR] 56-68) in the UK Biobank and 60 years (IQR 49-69) in the NHANES (**Table 1**; **Supplemental Tables 6 and 7**). The proportion of females was 55.7% (44,410) in the UK Biobank and 51.6% (2,432) in the NHANES. Current smokers constituted 6.9% (5,532) of the UK Biobank cohort and 17.8% (839) of the NHANES cohort. The prevalence of prior disease (i.e., myocardial infarction, stroke, congestive heart failure, cancer, or diabetes) was 13.6% (10,856) in the UK Biobank and 32.4% (1,528) in the NHANES.

**Table 1:** Baseline characteristics of the UK Biobank and the NHANES cohorts.

|  | UK Biobank<br>(N=79,717) | NHANES<br>(N=4,713) |
| --- | --- | --- |
| Female | 44,410 (55.7%) | 2,432 (51.6%) |
| Age at accelerometer wear (years) | 63 [56-68] | 60 [49-69] |
| <50 years | 4,713 (5.9%) | 1,195 (25.4%) |
| 50-54 years | 10,059 (12.6%) | 612 (13.0%) |
| 55-59 years | 11,843 (14.9%) | 543 (11.5%) |
| 60-64 years | 14,868 (18.7%) | 715 (15.2%) |
| 65-69 years | 20,020 (25.1%) | 528 (11.2%) |
| 70-74 years | 14,721 (18.5%) | 443 (9.4%) |
| ≥75 years | 3,493 (4.4%) | 677 (14.4%) |
| Current smoker | 5,532 (6.9%) | 839 (17.8%) |
| Body mass index (kg/m<sup>2</sup>) | 26.7 (4.5) | 29.5 (6.8) |
| Systolic blood pressure (mmHg) | 137 (18.1) | 128 (18.7) |
| Blood pressure medication | 13,689 (17.2%) | 1,895 (40.2%) |
| Cholesterol lowering medication | 11,343 (14.2%) | 1,437 (30.5%) |
| **Prior disease** |  |  |
| Myocardial infarction | 1,770 (2.2%) | 264 (5.6%) |
| Stroke | 997 (1.3%) | 237 (5.0%) |
| Congestive heart failure | 622 (0.8%) | 221 (4.7%) |
| Cancer | 5,583 (7.0%) | 480 (10.2%) |
| Diabetes | 3,276 (4.1%) | 860 (18.2%) |
| Any of the above prior diseases | 10,856 (13.6%) | 1,528 (32.4%) |
| **Accelerometer data** |  |  |
| Daily steps (steps per day) | 9,030 [6,790-11,700] | 7,780 [5,120-11,000] |
| Walking cadence (steps per minute) | 114 [106-121] | 103 [95-111] |

Data are number of participants (%), mean (standard deviation), and median [interquartile range]. Median accelerometer wear time was 7.0 days (interquartile range 6.8-7.0) in UK Biobank and 7.0 days (interquartile range 6.9-7.0) in NHANES. NHANES = National Health and Nutrition Examination Survey.

Median daily steps was 9,030 (IQR 6,790-11,700) in UK Biobank and 7,780 (IQR 5,120-11,000) in the NHANES, with median peak 1-min walking cadence of 114 steps per minute (IQR 106-121) and 103 steps per minute (IQR 95-111), respectively.

### All-cause mortality

In the UK Biobank, 2.1% (1,640) of participants died within the first five-years of follow-up. Adding accelerometer data to the baseline model (c-index 0.762, 95% confidence interval [CI] 0.752-0.774) resulted in modest but statistically significant improvements in discrimination for 5-year mortality (c-index 0.770, 95% CI 0.760-0.782; change in c-index 0.008, 95% CI 0.005-0.011; **Figure 1**; see **Supplemental Table 8** for further details about the developed model). In the NHANES, 7.9% (372) of participants died within the first five-years of follow-up, with similar improvements in the c-index observed (baseline model: c-index 0.801, 95% CI 0.778-0.820; baseline plus accelerometer data: 0.816, 0.795-0.835; change in c-index 0.015, 95% CI 0.007-0.025). Adding accelerometer data to the baseline model in the UK Biobank led to a 3.4% increase in the proportion of the 1,640 participants who died within 5-years being correctly classified as ≥10% risk, while only increasing the proportion of participants alive at 5-years being misclassified as high risk by <0.1%. Thus, the net reclassification index in the UK Biobank was 3.3% (95% CI 2.1%-4.5%; **Table 2**). Similarly, the net reclassification index in the NHANES was 4.0% (95% CI 0.7%-7.4%).

**Figure 1:**
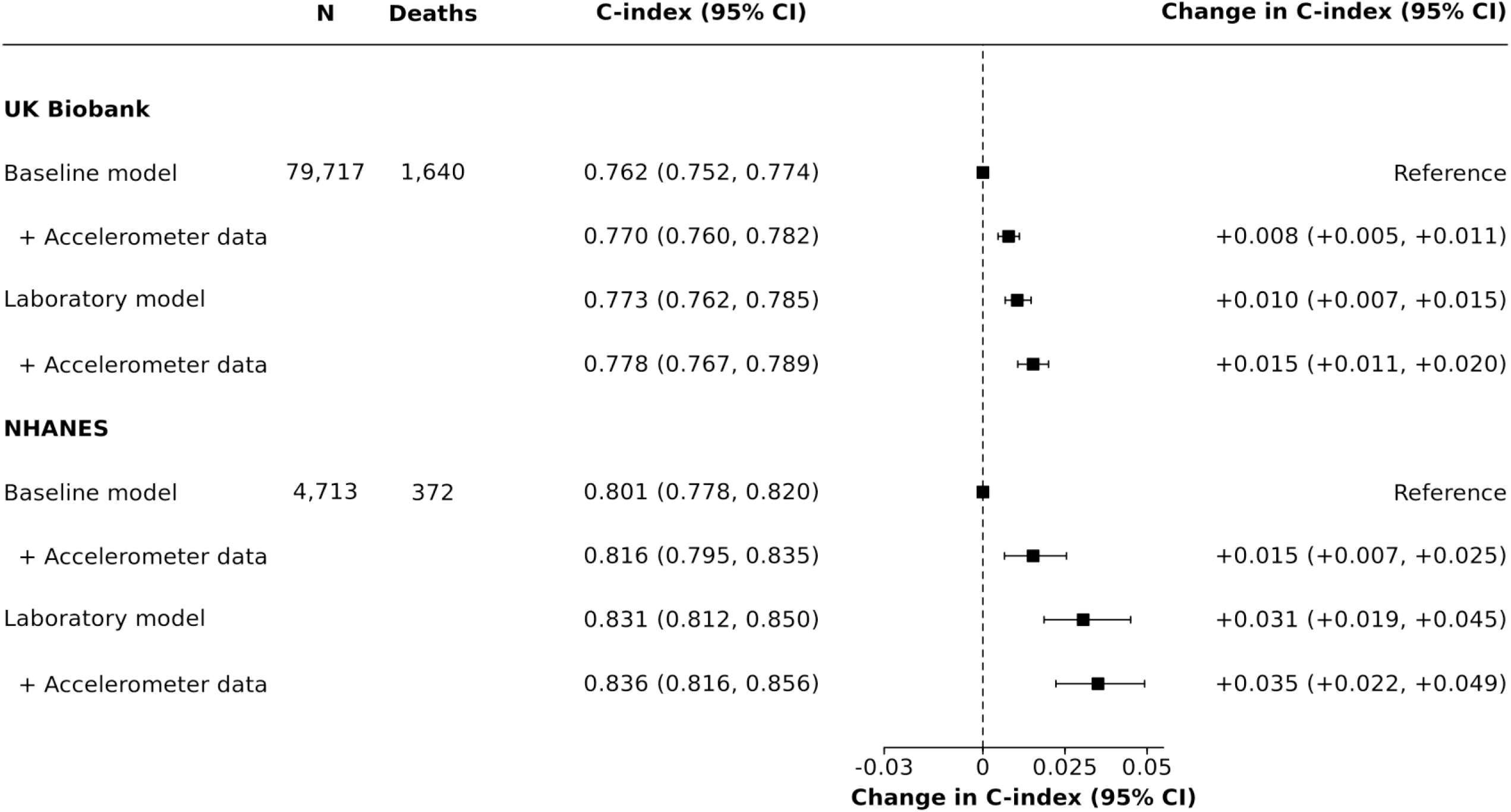
Discrimination improvement after addition of accelerometer data to baseline and laboratory models for 5-year predicted mortality risk Squares represent change in c-index compared to the baseline model and horizontal lines are 95% confidence intervals generated using 1,000 bootstrap samples. Baseline model: sex, age group, smoking, body mass index, systolic blood pressure, blood pressure medication, cholesterol medication, prior myocardial infarction, stroke, congestive heart failure, cancer, and diabetes. Laboratory model: baseline model plus albumin, alanine aminotransferase, aspartate aminotransferase, high density lipoprotein cholesterol, gamma glutamyltransferase, lymphocyte count, mean corpuscular haemoglobin, mean corpuscular haemoglobin concentration, red blood cell count, red blood cell distribution width, urea, and white blood cell count. Accelerometer data: daily steps and walking cadence. CI = Confidence interval. N = Number of participants. NHANES = National Health and Nutrition Examination Survey.

**Figure 2:**
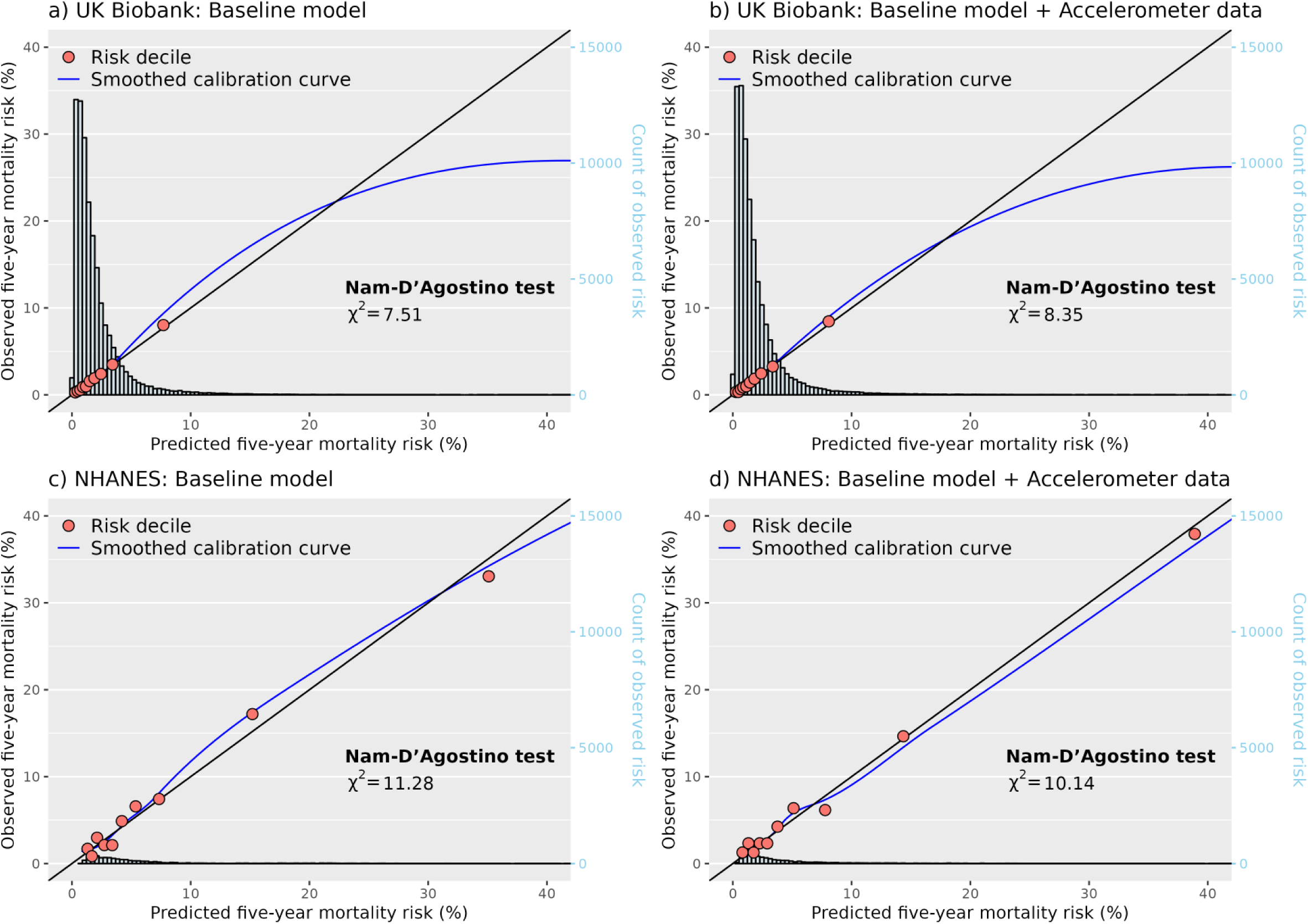
Calibration performance for baseline model and baseline model plus accelerometer data for 5-year all-cause mortality risk, by predicted risk decile Red circles represent the observed five-year mortality risk at each decile of predicted five-year risk. Black line represents perfect calibration. Blue line represents the smoothed calibration curve of all risk deciles generated using jack-knife methods. Data for calibration plots generated using five-fold cross validation. Calibration performance estimated using Nam-D’Agostino test with nine degrees of freedom, where the smaller values represents greater performance. Baseline model: sex, age group, smoking, body mass index, systolic blood pressure, blood pressure medication, cholesterol medication, prior myocardial infarction, stroke, congestive heart failure, cancer, and diabetes. Accelerometer data: daily steps and walking cadence. NHANES = National Health and Nutrition Examination Survey.

**Table 2:** Reclassification of participants after addition of accelerometer data to baseline and laboratory models for 5-year predicted mortality risk.

|  | UK Biobank (N=79,717) |  | NHANES (N=4,713) |  |
| --- | --- | --- | --- | --- |
|  | Baseline model versus baseline plus accelerometer data | Laboratory model versus laboratory plus accelerometer data | Baseline model versus baseline plus accelerometer data | Laboratory model versus laboratory plus accelerometer data |
| **Discrimination** |  |  |  |  |
| C-index change (95% CI) | 0.008 (0.005-0.011) | 0.005 (0.002-0.007) | 0.015 (0.007-0.025) | 0.005 (-0.001-0.011) |
| **Reclassification (10% risk threshold)** |  |  |  |  |
| Participants who died at 5-years |  |  |  |  |
| Appropriately reclassified* | 81 (4.9%) | 81 (4.9%) | 28 (7.5%) | 15 (4.0%) |
| Inappropriately reclassified | 25 (1.5%) | 29 (1.8%) | 11 (3.0%) | 14 (3.8%) |
| No change | 1,534 (93.5%) | 1,530 (93.3%) | 333 (89.5%) | 343 (92.2%) |
| Participants alive at 5-years |  |  |  |  |
| Appropriately reclassified** | 372 (0.5%) | 333 (0.4%) | 121 (2.8%) | 101 (2.3%) |
| Inappropriately reclassified | 446 (0.6%) | 376 (0.5%) | 144 (3.3%) | 85 (2.0%) |
| No change | 77,259 (99.0%) | 77,368 (99.1%) | 4,076 (93.9%) | 4,155 (95.7%) |
| Net reclassification index (95% CI) | 3.3% (2.1%-4.5%) | 3.1% (1.9%-4.4%) | 4.0% (0.7%-7.4%) | 0.6% (-2.3%-3.5%) |
| Integrated discrimination improvement (95% CI) | 0.006 (0.005-0.008) | 0.006 (0.004-0.007) | 0.050 (0.038-0.061) | 0.034 (0.024-0.044) |

Baseline model: sex, age, smoking, body mass index, systolic blood pressure, blood pressure medication, cholesterol medication, prior myocardial infarction, stroke, congestive heart failure, cancer, and diabetes. Laboratory model: baseline model plus albumin, alanine aminotransferase, aspartate aminotransferase, high density lipoprotein cholesterol, gamma glutamyltransferase, lymphocyte count, mean corpuscular haemoglobin, mean corpuscular haemoglobin concentration, red blood cell count, red blood cell distribution width, urea, and white blood cell count. Accelerometer data: daily steps and walking cadence. \*For participants who died at 5-years, appropriately reclassified refers to a change in predicted risk from below to above the 10% risk threshold (i.e. an increase in predicted risk). \*\*Similarly, for participants alive at 5-years, appropriately reclassified refers to a change in predicted risk from above to below the 10% risk threshold (i.e. a decrease in predicted risk). CI = Confidence interval. NHANES = National Health and Nutrition Examination Survey.

Improvements in prediction were somewhat attenuated when accelerometer data were added to the laboratory model. In the UK Biobank, where laboratory measures were collected a median of 5.7 years (IQR 4.9-6.5) before accelerometer wear, the laboratory model had a c-index of 0.773 (0.762-0.785), and adding accelerometer data only slightly improved discrimination (0.778, 0.767-0.789; change in c-index 0.005, 95% CI 0.002-0.007). In NHANES, where laboratory data was collected on the same day as accelerometer wear started, no improvement in discrimination was observed (change in c-index 0.005, -0.001-0.011). Reclassification performance showed similar levels of attenuation, with the UK Biobank and NHANES having net reclassification indexes of 3.1% (1.9%-4.4%) and 0.6% (-2.3%-3.5%), respectively. All four models were well calibrated in both the UK Biobank and the NHANES, with good performance for both low and high risk participants (Nam-D’Agostino χ^2^ range 6.8-13.2; **Figure 2**; **Supplemental Figure 3**).

### Subgroups

Adding accelerometer data to the baseline model improved discrimination across all subgroups in the UK Biobank cohort (**Supplemental Figures 4 and 5**). The largest increase in the c-index was observed in participants with prior disease at baseline (i.e., myocardial infarction, stroke, congestive heart failure, cancer, or diabetes), with a change in c-index of 0.028 (95% CI 0.019-0.039). Improvements were more modest for individuals with no prior disease, showing a change in c-index of 0.007 (95% CI 0.002- 0.012). Reclassification performance was also notably better for participants with prior disease, with a net reclassification index (NRI) of 5.9% (95% CI 3.1%-8.6%) compared to 1.8% (95% CI 0.1%-2.8%) for those without prior disease (**Supplemental Table 9**). Although the NHANES data had limited power to assess subgroup differences robustly, c-index point estimates were directionally consistent with the UK Biobank results. Specifically, the change in c-index for those with prior disease was 0.036 (95% CI 0.020-0.053), while for those without prior disease it was 0.007 (95% CI -0.013-0.031; **Supplemental Figures 6 and 7**).

### Cause of death

To determine which causes of death were better predicted with the inclusion of accelerometer data to the baseline model, we assessed the models’ discriminative performance separately for cardiovascular, cancer, and other causes of death. In the UK Biobank, no improvement in c-index was observed for vascular deaths (change in c-index 0.003, 95% CI -0.005-0.011) and only modesty for cancer deaths (change in c-index 0.004, 95% CI 0.001-0.009; **Supplemental Figure 8**). Improvements appeared to be solely driven by other causes of death, with a change in c-index of 0.023 (95% CI 0.015-0.034). Among the 322 deaths attributed to other causes, the most common were diseases of the respiratory system (84 deaths), diseases of the nervous system (52 deaths), diseases of the digestive system (52 deaths), and external causes of morbidity and mortality (40 deaths; **Supplemental Table 10**). Although NHANES had too few events to robustly test for differences between causes of death, point estimates were directionally consistent with those from the UK Biobank (**Supplemental Figure 9**; **Supplemental Table 11**).

Regression coefficients and model formulas for 5-year mortality risk scores are available in supplementary tables 12-13 for model translation and implantation. For example, the estimated 5-year mortality risk for a 60-year-old with risk factors current smoker, BMI of 27,160 mm Hg SBP, Cholesterol lowering medication, prior diabetes, 7000 steps per day using the full Laboratory model plus accelerometer model are shown in supplementary table 13.

## Discussion

This study, utilising data from the UK Biobank and externally validating the findings in the US National Health and Nutrition Examination Survey (NHANES), found that integrating 7 days of device-measured daily steps and walking cadence data modestly enhanced the prediction of five-year all-cause mortality risk beyond traditional risk factors. The improvements were consistent across various subgroups, with the most significant gains in predictive performance observed in individuals with prior disease (i.e., myocardial infarction, stroke, congestive heart failure, cancer, or diabetes). When analysing the impact of accelerometer data on different causes of death within the UK Biobank, we found no significant improvement in prediction for vascular deaths and only modest changes for cancer deaths. Instead, lower daily steps and slower walking cadence appeared to be more important predictors for deaths attributed to respiratory, nervous system, and digestive system diseases. Given the widespread use of digital devices capable of tracking daily step counts and walking cadence, these findings highlight their potential utility for public health organizations.^7^ By leveraging such data, it may be possible to remotely monitor large populations and better target interventions to individuals at higher risk, thereby enhancing public health strategies and outcomes.^6^

This study offers important advances in our understanding of how device-measured physical activity behaviours can improve prediction of future health outcomes. By focusing on two easily understood physical activity metrics reported by consumer-grade devices, daily step count and walking cadence, the results from this report are directly applicable to public health organisations. Through external validation, we demonstrated that these simple metrics provide comparable performance to the complex behaviours (e.g., relative amplitude and standard deviation of the 6th principal component) evaluated by previous studies, which reported changes in c-index ranging from 0.004 to 0.029 and area under the curve from 0.014 to 0.040.^8–13^ Furthermore, our novel findings highlight that the greatest improvements in prediction may be in individuals with prior disease, indicating that device-measured physical activity metrics are particularly valuable for identifying higher-risk individuals. The only previous study to investigate differences by subpopulations was Chen at al^8^ finding no differences between participants with one or more morbidity versus none. However, this study was substantially underpowered to investigate such differences, with only 410 (n=3,991) and 105 (n=1,329) deaths in the internal and external validation cohorts, respectively.

Our study has several strengths and limitations. It is the largest published study to-date investigating whether accelerometer data can enhance mortality prediction, with results validated externally. We employed an open-source algorithm to derive step count and walking cadence, which ensured transparency and supports the replication of our methods.^19^ In addition to assessing discrimination metrics, we evaluated model calibration and reclassification, providing a more comprehensive understanding of performance. The study’s substantial statistical power enabled us to explore critical aspects of mortality prediction, such as identifying which causes of death were most accurately predicted and examining differences across sub-populations. However, our analysis was limited to seven days of accelerometer wear. Despite this, previous research has shown repeated 7-day measurements have a good infraclass correlation coefficient (>0.6), suggesting that this is unlikely to be a major source of error.^16^ The substantial differences in mortality rates and baseline characteristics between the two cohorts required us to recalibrate all four models in NHANES, including updating the model coefficients and baseline hazard, which may have limited the extent of true external validation. On the other hand, this heterogeneity between cohorts allowed us to test our hypothesis in both low- and high-risk populations, offering a more robust overall assessment of its validity.

The present study demonstrated that device-measured daily step counts and walking cadence can modestly improve mortality risk prediction beyond traditional risk factors, with the most significant enhancements observed in higher-risk individuals with pre-existing conditions. Our findings highlight the potential of integrating physical activity data, already routinely recorded by digital devices owned by most adults in high-income countries, into existing risk models. By incorporating these metrics, public health strategies can be refined to more accurately assess and manage mortality risk, particularly in older populations.

## Research in context

### Evidence before the study

We conducted a search in MEDLINE on July 22 2024, using the terms (predict*) AND (mortality OR death) AND (acceleromete*) AND (“physical activity”), restricted to English language. This search identified seven relevant articles that examined the predictive performance of device-measured physical activity behaviours. These studies reported modest improvements in mortality prediction when accelerometer data were added to traditional risk models. However, they primarily focused on physical activity metrics not commonly reported by consumer-grade devices or easily understood by the general public, which limits their applicability in current public health decision-making.

### Added value of the study

Our research addressed this gap by focusing on two easily understood physical activity metrics reported by consumer-grade devices: daily step count and peak 1-minute walking cadence. By developing our prediction model using data from the UK Biobank and validating it externally with the US 2011-2014 National Health and Nutrition Examination Survey (NHANES), we found that these metrics modestly improved the prediction of five-year all-cause mortality beyond traditional risk factors. Furthermore, the greatest improvements in risk discrimination were observed in individuals with prior disease.

### Implications of all the available evidence

Current evidence suggests that device-measured physical activity behaviours are strongly associated with mortality risk and can enhance prediction when combined with traditional risk factors. These findings indicate the potential for using such data to remotely monitor large populations and more precisely target healthcare interventions to those individuals at higher risk, thereby improving public health strategies and outcomes.

## Authorship Contributions

AD: conceptualisation, data curation, formal analysis, methodology, supervision, writing - review & editing. ADS: data curation, formal analysis, methodology, validation, writing - review & editing. BM: data curation, formal analysis, methodology, writing - review & editing. CH: conceptualisation, formal analysis, methodology, visualisation, validation, writing - original draft. DB: conceptualisation, formal analysis, methodology, supervision, writing - review & editing. SC: data curation, formal analysis, methodology, writing - review & editing. DM, PP, JS, AS: methodology.

## Data Sharing

The data used in this analysis is available to approved researchers from the UK Biobank.

## Data Availability

All data used in this analysis is available to approved researchers from the UK Biobank.

## Acknowledgements

We would like to thank the UK Biobank team and participants, without whom this research would not be possible.

## Funder Acknowledgements

AD’s research team is supported by a range of grants from the Wellcome Trust [223100/Z/21/Z, 227093/Z/23/Z], Novo Nordisk, Swiss Re, Boehringer Ingelheim, National Institutes of Health’s Oxford Cambridge Scholars Program, EPSRC Centre for Doctoral Training in Health Data Science (EP/S02428X/1), British Heart Foundation Centre of Research Excellence (grant number RE/18/3/34214), and funding administered by the Danish National Research Foundation in support of the Pioneer Centre for SMARTbiomed. For the purpose of open access, the author(s) has applied a Creative Commons Attribution (CC BY) licence to any Author Accepted Manuscript version arising..

