## Supplementary Material for "Predictive performance of wearable sensors for mortality risk in older adults: a model development and validation study"

**Predictive performance of daily step count and walking cadence for mortality risk in older adults: a model development and validation study**

### Supplemental Materials


Supplemental Table 1: Power calculations for risk models in the UK Biobank and NHANES ... 2
Supplemental Table 2: Derivation of risk factors in the UK Biobank ... 3
Supplemental Table 3: Derivation of risk factors in the NHANES ... 4
Supplemental Table 4: Codes used to define prior diseases in the UK Biobank ... 5
Supplemental Table 5: TRIPOD Checklist ... 6
Supplemental Table 6: Baseline characteristics of the UK Biobank cohort, by sex ... 7
Supplemental Table 7: Baseline characteristics of the NHANES cohort, by sex ... 8
Supplemental Table 8: Adjusted hazard ratios for predictors in the UK Biobank, by risk prediction model ... 9
Supplemental Table 9: Reclassification of participants after addition of accelerometer data to baseline and laboratory models for 5-year predicted mortality risk, by subgroup ... 10
Supplemental Table 10: Underlying cause of death at five-years in the UK Biobank ... 11
Supplemental Table 11: Underlying cause of death at five-years in the NHANES ... 12
Supplemental Table 12: 5-year mortality risk equations ... 13
Supplemental Table 13: Example 5-year mortality risk score of 65-year-old male ... 16
Supplemental Figure 1: Flow chart for UK Biobank participants included in analyses ... 18
Supplemental Figure 2: Flow chart for NHANES participants included in analyses ... 19
Supplemental Figure 3: Calibration of laboratory models for 5-year all-cause mortality risk in the UK Biobank and the NHANES, by predicted risk decile ... 20
Supplemental Figure 4: Discrimination improvement after addition of accelerometer data to baseline model for 5-year predicted mortality risk in the UK Biobank, by subgroup ... 21
Supplemental Figure 5: Discrimination improvement after addition of accelerometer data to laboratory model for 5-year predicted mortality risk in the UK Biobank, by subgroup ... 22
Supplemental Figure 6: Discrimination improvement after addition of accelerometer data to baseline model for 5-year predicted mortality risk in the NHANES, by subgroup ... 23
Supplemental Figure 7: Discrimination improvement after addition of accelerometer data to laboratory model for 5-year predicted mortality risk in the NHANES, by subgroup ... 24
Supplemental Figure 8: Discrimination improvement after addition of accelerometer data to baseline model for 5-year predicted mortality risk, by cause of death ... 25
Supplemental Figure 9: Discrimination improvement after addition of accelerometer data to laboratory model for 5-year predicted mortality risk, by cause of death ... 26

### Supplemental Table 1: Power calculations for risk models in the UK Biobank and NHANES

| Cohort | Model | Number of parameters | Prevalence of 5-year mortality | C-statistic | Estimated number of participants required | Estimated number of events required |
|---|---|---|---|---|---|---|
| UK Biobank | Baseline | 20 | 2.1% | 0.75 | 9,956 | 210 |
| UK Biobank | Laboratory | 40 | 2.1% | 0.75 | 19,912 | 419 |
| NHANES | Baseline | 20 | 7.8% | 0.80 | 1,875 | 147 |
| NHANES | Laboratory | 40 | 7.8% | 0.80 | 3,749 | 293 |

Power calculations conducted using the *pmsampsize* R package (version 1.1.3) based on Riley et al. *Statistics in Medicine*. 2019; 38: 1276-1296. NHANES = National Health and Nutrition Examination Survey.

### Supplemental Table 2: Derivation of risk factors in the UK Biobank

| Risk factor | Units | UKB field ID | Fractional polynomial |
|---|---|---|---|
| Age at accelerometer wear | 5-year age groups† | 90011; 34; 52 | |
| Sex | Male; Female | 31 | |
| Current smoker | Yes; Other | 20116 | |
| Body mass index | Kilograms per metre$^2$ | 21001 | |
| Systolic blood pressure | Millimetres of mercury | 93 | |
| Blood pressure medication | Yes; Other | 6177; 6153 | |
| Cholesterol lowering medication | Yes; Other | 6177; 6153 | |
| **Prior disease*** | | | |
| Myocardial infarction | Yes; No | 20002 | |
| Stroke | Yes; No | 20002 | |
| Congestive heart failure | Yes; No | 20002 | |
| Cancer | Yes; No | | |
| Diabetes | Yes; No | 20002 | |
| **Laboratory measures** | | | |
| Albumin | Grams per litre | 30600 | |
| Alanine aminotransferase | Microliter | 30620 | |
| Aspartate aminotransferase | Microliter | 30650 | $AST^2 + \log(AST)AST^2$ |
| HDL cholesterol | Millimoles per litre | 30760 | |
| Gamma glutamyltransferase | Microliter | 30730 | |
| Lymphocyte count | 10^9 cells per litre | 30120 | $Lymphocyte^{-0.5}$ |
| Mean corpuscular haemoglobin | Petagram | 30050 | |
| Mean corpuscular haemoglobin concentration | Grams per decilitre | 30060 | $MCHC^3$ |
| Red blood cell count | 10^12 cells per litre | 30010 | $RBC^3 + \log(RBC)RBC^3$ |
| Red blood cell distribution width | Percentage | 30070 | |
| Urea | Millimoles per litre | 30670 | $Urea^{-0.5} + \log(Urea)$ |
| White blood cell count | 10^9 cells per litre | 30000 | $WBC^2 + WBC^3$ |
| **Accelerometer data**** | | | |
| Daily steps | Steps per day | | Steps * log (Steps) |
| Walking cadence | Steps per minute | | |

†Age groups include <50, 50-54, 55-59, 60-64, 65-69, 70-74, and ≥75 years. *Derived from self-reported and electronic health record data, except cancer where only electronic health records were used. **Measures derived from raw accelerometer data (UKB field ID: 90001). HDL = High density lipoprotein. ID = Identifier. MCHC = Mean corpuscular haemoglobin concentration. RBC = Red blood count. WBC = White blood count.

### Supplemental Table 3: Derivation of risk factors in the NHANES

| Risk factor | Units | NHANES Field ID | Fractional polynomial |
|---|---|---|---|
| Age at accelerometer wear | 5-year age groups† | RIDAGEYR | |
| Sex | Male; Female | RIAGENDR | |
| Current smoker | Yes; Other | SMQ040 | |
| Body mass index | Kilograms per $metre^2$ | BMXBMI | |
| Systolic blood pressure | Millimetres of mercury | BPXSY1-BPXSY3 | |
| Blood pressure medication | Yes; No | BPQ050A | |
| Cholesterol lowering medication | Yes; No | BPQ100D | |
| **Prior disease*** | | | |
| Myocardial infarction | Yes; No | MCQ160E | |
| Stroke | Yes; No | MCQ160F | |
| Congestive heart failure | Yes; No | MCQ160B | |
| Cancer | Yes; No | MCQ220 | |
| Diabetes | Yes; No | DIQ010 | |
| **Laboratory measures** | | | |
| Albumin | Grams per litre | LBDSALSI | |
| Alanine aminotransferase | Microliter | LBXSATSI | |
| Aspartate aminotransferase | Microliter | LBXSASSI | $AST^2 + log(AST)AST^2$ |
| HDL cholesterol | Millimoles per litre | LBDHDDSI | |
| Gamma glutamyltransferase | Microliter | LBXSGTSI | |
| Lymphocyte count | 10^9 cells per litre | LBDLYMNO | $Lymphocyte^{-0.5}$ |
| Mean corpuscular haemoglobin | Petagrams | LBXMCHSI | |
| Mean corpuscular haemoglobin concentration | Grams per decilitre | LBXMC | $MCHC^3$ |
| Red blood cell count | 10^12 cells per litre | LBXRBCSI | $RBC^3 + log(RBC)RBC^3$ |
| Red blood cell distribution width | Percentage | LBXRDW | |
| Urea | Millimoles per litre | LBDSBUSI | $Urea^{-0.5} + log(Urea)$ |
| White blood cell count | 10^9 cells per litre | LBXRDW | $WBC^2 + WBC^3$ |
| **Accelerometer data**** | | | |
| Daily steps | Steps per day | | Steps * log(Steps) |
| Walking cadence | Steps per minute | | |

†Age groups include <50, 50-54, 55-59, 60-64, 65-69, 70-74, and ≥75 years. *Derived from self-reported data only. **Measures derived from raw accelerometer data (NHANES field ID: PAX80). HDL = High density lipoprotein. ID = Identifier. MCHC = Mean corpuscular haemoglobin concentration. NHANES = National Health and Nutrition Examination Survey. RBC = Red blood count. WBC = White blood count.

### Supplemental Table 4: Codes used to define prior diseases in the UK Biobank

| Prior disease | ICD-10 diagnosis codes in EHR data | Self-reported conditions at baseline |
|---|---|---|
| Myocardial infarction | I21; I22; I25.2 | “heart attack/myocardial infarction” |
| Stroke | I60-I64 | “stroke” |
| Heart failure | I11.0; I13.0; I13.2; I25.5; I42-I43; I50; P29.0 | “heart failure/pulmonary oedema” |
| Cancer | C00-C97 (excluding C44 non-melanoma skin cancer)† | |
| Diabetes | E10-E14 | “diabetes”; ”type 1 diabetes”; ”type 2 diabetes” |

Prior disease in UK Biobank were identified using electronic health record data, where an ICD-10 code was present prior to accelerometer wear (with all participants having a 10-year look back period), or reported by the participant at their baseline assessment (UK Biobank field ID: 20002). †Prior cancer identified in UK Biobank using routinely collected hospital admission and cancer registry data, and excludes non-melanoma skin cancer (ICD-10 code C44). EHR = Electronic Health Record. ICD-10 = International Classification of Diseases, 10$^{th}$ revision.

### Supplemental Table 5: TRIPOD Checklist

| Section/Topic | | | Checklist Item | Page |
|---|---|---|---|---|
| **Title and abstract** | | | | |
| Title | 1 | D;V | Identify the study as developing and/or validating a multivariable prediction model, the target population, and the outcome to be predicted. | 1 |
| Abstract | 2 | D;V | Provide a summary of objectives, study design, setting, participants, sample size, predictors, outcome, statistical analysis, results, and conclusions. | 2 |
| **Introduction** | | | | |
| Background and objectives | 3a | D;V | Explain the medical context (including whether diagnostic or prognostic) and rationale for developing or validating the multivariable prediction model, including references to existing models. | 3 |
| | 3b | D;V | Specify the objectives, including whether the study describes the development or validation of the model or both. | |
| **Methods** | | | | |
| Source of data | 4a | D;V | Describe the study design or source of data (e.g., randomized trial, cohort, or registry data), separately for the development and validation data sets, if applicable. | 4 |
| | 4b | D;V | Specify the key study dates, including start of accrual; end of accrual; and, if applicable, end of follow-up. | 4 |
| Participants | 5a | D;V | Specify key elements of the study setting (e.g., primary care, secondary care, general population) including number and location of centres. | 4 |
| | 5b | D;V | Describe eligibility criteria for participants. | 5 |
| | 5c | D;V | Give details of treatments received, if relevant. | X |
| Outcome | 6a | D;V | Clearly define the outcome that is predicted by the prediction model, including how and when assessed. | 5 |
| | 6b | D;V | Report any actions to blind assessment of the outcome to be predicted. | X |
| Predictors | 7a | D;V | Clearly define all predictors used in developing or validating the multivariable prediction model, including how and when they were measured. | 4-6 |
| | 7b | D;V | Report any actions to blind assessment of predictors for the outcome and other predictors. | X |
| Sample size | 8 | D;V | Explain how the study size was arrived at. | 7 |
| Missing data | 9 | D;V | Describe how missing data were handled (e.g., complete-case analysis, single imputation, multiple imputation) with details of any imputation method. | 5 |
| Statistical analysis methods | 10a | D | Describe how predictors were handled in the analyses. | 6 |
| | 10b | D | Specify type of model, all model-building procedures (including any predictor selection), and method for internal validation. | 5-6 |
| | 10c | V | For validation, describe how the predictions were calculated. | 6 |
| | 10d | D;V | Specify all measures used to assess model performance and, if relevant, to compare multiple models. | 6-7 |
| | 10e | V | Describe any model updating (e.g., recalibration) arising from the validation, if done. | 6 |
| Risk groups | 11 | D;V | Provide details on how risk groups were created, if done. | 6 |
| Development vs. validation | 12 | V | For validation, identify any differences from the development data in setting, eligibility criteria, outcome, and predictors. | 6 |
| **Results** | | | | |
| Participants | 13a | D;V | Describe the flow of participants through the study, including the number of participants with and without the outcome and, if applicable, a summary of the follow-up time. A diagram may be helpful. | 7-8 |
| | 13b | D;V | Describe the characteristics of the participants (basic demographics, clinical features, available predictors), including the number of participants with missing data for predictors and outcome. | 7 |
| | 13c | V | For validation, show a comparison with the development data of the distribution of important variables (demographics, predictors and outcome). | 7-8 |
| Model development | 14a | D | Specify the number of participants and outcome events in each analysis. | 7 |
| | 14b | D | If done, report the unadjusted association between each candidate predictor and outcome. | X |
| Model specification | 15a | D | Present the full prediction model to allow predictions for individuals (i.e., all regression coefficients, and model intercept or baseline survival at a given time point). | 8 |
| | 15b | D | Explain how to the use the prediction model. | |
| Model performance | 16 | D;V | Report performance measures (with CIs) for the prediction model. | 8 |
| Model-updating | 17 | V | If done, report the results from any model updating (i.e., model specification, model performance). | 8 |
| **Discussion** | | | | |
| Limitations | 18 | D;V | Discuss any limitations of the study (such as nonrepresentative sample, few events per predictor, missing data). | 10-11 |
| Interpretation | 19a | V | For validation, discuss the results with reference to performance in the development data, and any other validation data. | 10-11 |
| | 19b | D;V | Give an overall interpretation of the results, considering objectives, limitations, results from similar studies, and other relevant evidence. | 11 |
| Implications | 20 | D;V | Discuss the potential clinical use of the model and implications for future research. | 11 |
| **Other information** | | | | |
| Supplementary information | 21 | D;V | Provide information about the availability of supplementary resources, such as study protocol, Web calculator, and data sets. | 13 |
| Funding | 22 | D;V | Give the source of funding and the role of the funders for the present study. | 13 |

### Supplemental Table 6: Baseline characteristics of the UK Biobank cohort, by sex

| | **Overall (N=79,717)** | **Female (N=44,410)** | **Male (N=35,307)** |
|---|---|---|---|
| Age at accelerometer wear (years) | | | |
| <50 years | 4,713 (5.9%) | 2,741 (6.2%) | 1,972 (5.6%) |
| 50-54 years | 10,059 (12.6%) | 5,983 (13.5%) | 4,076 (11.5%) |
| 55-59 years | 11,843 (14.9%) | 7,123 (16.0%) | 4,720 (13.4%) |
| 60-64 years | 14,868 (18.7%) | 8,711 (19.6%) | 6,157 (17.4%) |
| 65-69 years | 20,020 (25.1%) | 10,879 (24.5%) | 9,141 (25.9%) |
| 70-74 years | 14,721 (18.5%) | 7,346 (16.5%) | 7,375 (20.9%) |
| ≥75 years | 3,493 (4.4%) | 1,627 (3.7%) | 1,866 (5.3%) |
| Current smoker | 5,532 (6.9%) | 2,603 (5.9%) | 2,929 (8.3%) |
| Body mass index (kg/m$^2$) | 26.7 (4.5) | 26.2 (4.8) | 27.3 (4.0) |
| Systolic blood pressure (mmHg) | 137 (18.1) | 134 (18.5) | 140 (16.9) |
| Blood pressure medication | 13,689 (17.2%) | 6,036 (13.6%) | 7,653 (21.7%) |
| Cholesterol lowering medication | 11,343 (14.2%) | 4,126 (9.3%) | 7,217 (20.4%) |
| **Prior disease** | | | |
| Myocardial infarction | 1,770 (2.2%) | 360 (0.8%) | 1,410 (4.0%) |
| Stroke | 997 (1.3%) | 392 (0.9%) | 605 (1.7%) |
| Congestive heart failure | 622 (0.8%) | 162 (0.4%) | 460 (1.3%) |
| Cancer | 5,583 (7.0%) | 3,068 (6.9%) | 2,515 (7.1%) |
| Diabetes | 3,276 (4.1%) | 1,223 (2.8%) | 2,053 (5.8%) |
| Any of the above prior diseases | 10,856 (13.6%) | 4,827 (10.9%) | 6,029 (17.1%) |
| **Laboratory data** | | | |
| Albumin (g/L) | 45.4 (2.6) | 45.1 (2.6) | 45.7 (2.6) |
| Alanine aminotransferase (U/L) | 22.7 (13.3) | 19.6 (11.4) | 26.6 (14.4) |
| Aspartate aminotransferase (U/L) | 25.8 (9.3) | 24.2 (8.6) | 27.8 (9.8) |
| HDL cholesterol (mmol/L) | 1.5 (0.4) | 1.6 (0.4) | 1.3 (0.3) |
| Gamma glutamyltransferase (U/L) | 34.2 (35.4) | 27.9 (29.1) | 42.0 (40.7) |
| Lymphocyte count (10^9 cells/L) | 1.9 (0.9) | 2.0 (1.0) | 1.9 (0.9) |
| Mean corpuscular haemoglobin (pg) | 31.5 (1.8) | 31.3 (1.9) | 31.7 (1.7) |
| Mean corpuscular haemoglobin concentration (g/dL) | 34.5 (1.1) | 34.4 (1.1) | 34.6 (1.0) |
| Red blood cell count (10^12 cells/L) | 4.5 (0.4) | 4.3 (0.3) | 4.8 (0.4) |
| Red blood cell distribution width (%) | 13.4 (0.9) | 13.5 (1.0) | 13.4 (0.8) |
| Urea (mmol/L) | 5.3 (1.3) | 5.1 (1.3) | 5.6 (1.3) |
| White blood cell count (10^9 cells/L) | 6.7 (1.8) | 6.7 (1.9) | 6.7 (1.8) |
| **Accelerometer data** | | | |
| Daily steps (steps per day) | 9,030 [6,790-11,700] | 8,970 [6,770-11,500] | 9,110 [6,830-11,800] |
| Walking cadence (steps per minute) | 114 [106-121] | 116 [109-123] | 111 [104-117] |

Data are number of participants (%), mean (standard deviation) and median [interquartile range]. HDL = High-density lipoprotein.

### Supplemental Table 7: Baseline characteristics of the NHANES cohort, by sex

| | **Overall (N=4,713)** | **Female (N=2,432)** | **Male (N=2,281)** |
|---|---|---|---|
| Age at accelerometer wear (years) | | | |
| <50 years | 1,195 (25.4%) | 633 (26.0%) | 562 (24.6%) |
| 50-54 years | 612 (13.0%) | 330 (13.6%) | 282 (12.4%) |
| 55-59 years | 543 (11.5%) | 281 (11.6%) | 262 (11.5%) |
| 60-64 years | 715 (15.2%) | 357 (14.7%) | 358 (15.7%) |
| 65-69 years | 528 (11.2%) | 267 (11.0%) | 261 (11.4%) |
| 70-74 years | 443 (9.4%) | 236 (9.7%) | 207 (9.1%) |
| ≥75 years | 677 (14.4%) | 328 (13.5%) | 349 (15.3%) |
| Current smoker | 839 (17.8%) | 361 (14.8%) | 478 (21.0%) |
| Body mass index (kg/m$^2$) | 29.5 (6.8) | 30.0 (7.5) | 29.0 (5.9) |
| Systolic blood pressure (mmHg) | 128 (18.7) | 127 (19.7) | 128 (17.5) |
| Blood pressure medication | 1,895 (40.2%) | 1001 (41.2%) | 894 (39.2%) |
| Cholesterol lowering medication | 1,437 (30.5%) | 711 (29.2%) | 726 (31.8%) |
| **Prior disease** | | | |
| Myocardial infarction | 264 (5.6%) | 96 (3.9%) | 168 (7.4%) |
| Stroke | 237 (5.0%) | 132 (5.4%) | 105 (4.6%) |
| Congestive heart failure | 221 (4.7%) | 106 (4.4%) | 115 (5.0%) |
| Cancer | 480 (10.2%) | 265 (10.9%) | 215 (9.4%) |
| Diabetes | 860 (18.2%) | 412 (16.9%) | 448 (19.6%) |
| Any of the above prior diseases | 1,528 (32.4%) | 751 (30.9%) | 777 (34.1%) |
| **Laboratory data** | | | |
| Albumin (g/L) | 42.2 (3.1) | 41.6 (3.0) | 42.7 (3.0) |
| Alanine aminotransferase (U/L) | 24.3 (18.2) | 21.5 (12.9) | 27.2 (22.2) |
| Aspartate aminotransferase (U/L) | 25.7 (14.8) | 24.3 (12.8) | 27.2 (16.6) |
| HDL cholesterol (mmol/L) | 1.4 (0.4) | 1.5 (0.4) | 1.2 (0.4) |
| Gamma glutamyltransferase (U/L) | 29.5 (38.1) | 26.4 (32.9) | 32.8 (42.8) |
| Lymphocyte count (10^9 cells/Litre) | 2.1 (1.2) | 2.1 (1.2) | 2.0 (1.2) |
| Mean corpuscular haemoglobin (pg) | 30.5 (2.5) | 30.2 (2.6) | 30.8 (2.3) |
| Mean corpuscular haemoglobin concentration (g/dL) | 33.9 (1.1) | 33.8 (1.1) | 34.0 (1.0) |
| Red blood cell count (10^12 cells/L) | 4.6 (0.5) | 4.4 (0.4) | 4.8 (0.5) |
| Red blood cell distribution width (%) | 13.5 (1.4) | 13.6 (1.5) | 13.4 (1.2) |
| Urea (mmol/L) | 5.1 (2.3) | 4.9 (2.2) | 5.3 (2.4) |
| White blood cell count (10^9 cells/L) | 7.0 (2.3) | 7.0 (2.3) | 7.0 (2.3) |
| **Accelerometer data** | | | |
| Daily steps (steps per day) | 7,780 [5,120-11,000] | 7,270 [4,770-10,200] | 8,460 [5,520-12,000] |
| Walking cadence (steps per min) | 103 [95-111] | 104 [94-112] | 103 [95-110] |

Data are number of participants (%), mean (standard deviation) and median [interquartile range]. HDL = High-density lipoprotein. NHANES = National Health and Nutrition Examination Survey.

### Supplemental Table 8: Adjusted hazard ratios for predictors in the UK Biobank, by risk prediction model

| | **Baseline model** | **Baseline model plus accelerometer data** | **Laboratory model** | **Laboratory model plus accelerometer data** |
|---|---|---|---|---|
| Male | 1.49 (1.34-1.65) | 1.53 (1.38-1.69) | 1.43 (1.26-1.64) | 1.49 (1.30-1.70) |
| Age group | | | | |
| <50 years | Reference | Reference | Reference | Reference |
| 50-54 years | 1.53 (0.87-2.67) | 1.52 (0.87-2.66) | 1.51 (0.87-2.64) | 1.51 (0.86-2.64) |
| 55-59 years | 2.38 (1.40-4.02) | 2.36 (1.39-3.99) | 2.31 (1.36-3.91) | 2.28 (1.35-3.86) |
| 60-64 years | 3.53 (2.12-5.87) | 3.44 (2.07-5.73) | 3.46 (2.08-5.77) | 3.34 (2.00-5.57) |
| 65-69 years | 5.02 (3.04-8.29) | 4.81 (2.92-7.95) | 4.88 (2.95-8.07) | 4.61 (2.79-7.63) |
| 70-74 years | 7.97 (4.83-13.16) | 7.28 (4.41-12.03) | 7.39 (4.46-12.24) | 6.70 (4.04-11.10) |
| ≥75 years | 11.11 (6.64-18.57) | 9.48 (5.66-15.88) | 9.83 (5.85-16.51) | 8.41 (5.00-14.15) |
| Current smoker | 2.27 (1.97-2.63) | 2.00 (1.72-2.31) | 1.89 (1.62-2.20) | 1.71 (1.46-1.99) |
| Body mass index (per 5 kg/m$^2$) | 1.16 (1.10-1.23) | 1.05 (0.99-1.11) | 1.13 (1.06-1.20) | 1.04 (0.98-1.10) |
| Systolic blood pressure (per 10 mmHg) | 1.08 (1.05-1.10) | 1.08 (1.05-1.11) | 1.09 (1.06-1.12) | 1.09 (1.06-1.12) |
| Blood pressure medication | 1.15 (1.01-1.30) | 1.10 (0.97-1.25) | 1.07 (0.95-1.22) | 1.04 (0.92-1.18) |
| Cholesterol lowering medication | 1.04 (0.91-1.19) | 1.04 (0.91-1.19) | 1.05 (0.92-1.20) | 1.05 (0.91-1.20) |
| **Prior disease** | | | | |
| Myocardial infarction | 1.51 (1.23-1.86) | 1.45 (1.18-1.77) | 1.42 (1.16-1.74) | 1.38 (1.12-1.69) |
| Stroke | 1.32 (0.99-1.76) | 1.16 (0.87-1.54) | 1.21 (0.91-1.61) | 1.09 (0.82-1.46) |
| Congestive heart failure | 2.34 (1.80-3.03) | 1.99 (1.54-2.59) | 2.09 (1.61-2.71) | 1.85 (1.42-2.40) |
| Cancer | 3.07 (2.73-3.44) | 2.95 (2.62-3.31) | 2.91 (2.59-3.27) | 2.83 (2.51-3.18) |
| Diabetes | 1.52 (1.28-1.80) | 1.38 (1.16-1.64) | 1.44 (1.21-1.72) | 1.34 (1.13-1.60) |
| **Laboratory measures** | | | | |
| Albumin (per 10 g/L) | | | 0.65 (0.53-0.79) | 0.67 (0.55-0.81) |
| ALT (per 10 U/L) | | | 0.92 (0.86-0.97) | 0.92 (0.87-0.98) |
| AST (per 10 U/L) FP1 | | | 1.07 (1.03-1.10) | 1.07 (1.04-1.10) |
| AST (per 10 U/L) FP2 | | | 0.98 (0.97-0.99) | 0.98 (0.97-0.99) |
| HDL cholesterol (per mmol/L) | | | 0.88 (0.75-1.03) | 0.93 (0.79-1.08) |
| Gamma glutamyltransferase (per 100 U/L) | | | 1.14 (1.04-1.25) | 1.10 (1.01-1.20) |
| Lymphocyte count (per 10^9 cells/Litre) | | | 1.60 (1.06-2.39) | 1.47 (0.98-2.21) |
| Mean corpuscular haemoglobin (per 10 pg) | | | 2.34 (1.61-3.40) | 2.21 (1.52-3.21) |
| Mean corpuscular haemoglobin concentration (per 10 g/dL) | | | 0.97 (0.96-0.99) | 0.97 (0.96-0.99) |
| Red blood count (per 10^12 cells/L) FP1 | | | 0.96 (0.93-0.99) | 0.96 (0.93-0.99) |
| Red blood count (per 10^12 cells/L) FP2 | | | 1.02 (1.01-1.04) | 1.02 (1.01-1.04) |
| Red blood distribution width (per 10%) | | | 3.78 (2.32-6.16) | 3.29 (1.99-5.45) |
| Urea (per 100 mmol/L) FP1 | | | 4.48 (2.48-8.09) | 3.47 (1.89-6.37) |
| Urea (per 100 mmol/L) FP2 | | | 23.06 (6.53-81.43) | 13.91 (3.81-50.78) |
| White blood count (per 10^10 cells/L) FP1 | | | 2.48 (1.75-3.52) | 2.11 (1.50-2.97) |
| White blood count (per 10^10 cells/L) FP2 | | | 0.77 (0.66-0.90) | 0.81 (0.70-0.94) |
| **Accelerometer data** | | | | |
| Daily steps (per 1000 steps) FP1 | | 0.64 (0.57-0.71) | | 0.68 (0.60-0.76) |
| Daily steps (per 1000 steps) FP2 | | 1.13 (1.09-1.17) | | 1.11 (1.07-1.15) |
| Walking cadence (per 10 steps/minute) | | 0.92 (0.88-0.97) | | 0.93 (0.89-0.98) |
| **Baseline survival estimate at 5 years** | 0.998 | 0.997 | 0.997 | 0.997 |

Data are adjusted hazard ratios with 95% confidence intervals in parenthesis. ALT = Alanine aminotransferase. AST = Aspartate aminotransferase. FP = Fractional polynomial. HDL = High-density lipoprotein.

### Supplemental Table 9: Reclassification of participants after addition of accelerometer data to baseline and laboratory models for 5-year predicted mortality risk, by subgroup

| | UK Biobank (N=79,717) | | NHANES (N=4,713) | |
|---|---|---|---|---|
| | **Baseline model versus baseline plus accelerometer data** | **Laboratory model versus laboratory plus accelerometer data** | **Baseline model versus baseline plus accelerometer data** | **Laboratory model versus laboratory plus accelerometer data** |
| **Net reclassification index (95% CI)** | | | | |
| Female | 3.1% (1.8%-4.5%) | 1.1% (-0.0%-2.5%) | 4.1% (-0.7%-8.9%) | 3.8% (-0.9%-8.4%) |
| Male | 3.5% (1.5%-5.3%) | 4.4% (2.6%-6.3%) | 3.7% (-1.0%-8.4%) | -2.0% (-5.7%-1.7%) |
| <70 years | 3.3% (2.0%-4.6%) | 2.7% (1.4%-3.9%) | 8.9% (2.4%-15.4%) | -0.0% (-5.5%-5.4%) |
| ≥70 years | 3.3% (1.0%-5.7%) | 3.7% (1.3%-6.1%) | 5.9% (1.6%-10.2%) | 5.0% (1.1%-8.9%) |
| Prior disease | 5.9% (3.1%-8.6%) | 5.1% (2.5%-7.7%) | 6.0% (1.6%-10.5%) | 1.0% (-2.8%-4.8%) |
| No prior disease | 1.8% (0.1%-2.8%) | 2.0% (0.8%-3.2%) | 1.4% (-4.2%-7.0%) | -0.3% (-5.1%-4.5%) |
| **Integrated discrimination improvement (95% CI)** | | | | |
| Female | 0.005 (0.003-0.007) | 0.005 (0.003-0.007) | 0.051 (0.034-0.069) | 0.036 (0.022-0.051) |
| Male | 0.007 (0.005-0.009) | 0.006 (0.004-0.008) | 0.049 (0.032-0.065) | 0.032 (0.019-0.046) |
| <70 years | 0.005 (0.004-0.007) | 0.005 (0.003-0.007) | 0.018 (0.008-0.029) | 0.018 (0.008-0.027) |
| ≥70 years | 0.008 (0.005-0.011) | 0.006 (0.004-0.009) | 0.077 (0.059-0.095) | 0.049 (0.035-0.064) |
| Prior disease | 0.011 (0.008-0.015) | 0.010 (0.006-0.013) | 0.063 (0.046-0.079) | 0.043 (0.030-0.057) |
| No prior disease | 0.003 (0.002-0.004) | 0.003 (0.002-0.004) | 0.031 (0.016-0.047) | 0.019 (0.006-0.032) |

Net reclassification index based on 10% risk threshold. Baseline model: sex, age group, smoking, body mass index, systolic blood pressure, blood pressure medication, cholesterol medication, prior myocardial infarction, stroke, congestive heart failure, cancer, and diabetes. Laboratory model: baseline model plus albumin, alanine aminotransferase, aspartate aminotransferase, high density lipoprotein cholesterol, gamma glutamyltransferase, lymphocyte count, mean corpuscular haemoglobin, mean corpuscular haemoglobin concentration, red blood cell count, red blood cell distribution width, urea, and white blood cell count. Accelerometer data: daily steps and walking cadence. CI = Confidence interval. N = Number of participants. NHANES = National Health and Nutrition Examination Survey.

### Supplemental Table 10: Underlying cause of death at five-years in the UK Biobank

| **Underlying cause of death (ICD-10)** | **N (%)** |
|---|---|
| Vascular death (I00-I99) | 334 (20.4%) |
| Cancer death (C00-C97) | 984 (60.0%) |
| Other death | 322 (19.6%) |
| Certain infectious and parasitic diseases (A) | 9 (0.5%) |
| Diseases of the blood and blood-forming organs and certain disorders involving the immune mechanism (D) | 23 (1.4%) |
| Endocrine, nutritional and metabolic diseases (E) | 11 (0.7%) |
| Mental and behavioural disorders (F) | 7 (0.4%) |
| Diseases of the nervous system (G) | 52 (3.2%) |
| Diseases of the ear and mastoid process (H) | 1 (0.1%) |
| Diseases of the respiratory system (J) | 84 (5.1%) |
| Diseases of the digestive system (K) | 52 (3.2%) |
| Diseases of the skin and subcutaneous tissue (L) | 4 (0.2%) |
| Diseases of the musculoskeletal system and connective tissue (M) | 9 (0.5%) |
| Diseases of the genitourinary system (N) | 6 (0.4%) |
| Congenital malformations, deformations and chromosomal abnormalities (Q) | 2 (0.1%) |
| Symptoms, signs and abnormal clinical and laboratory findings, not elsewhere classified (R) | 6 (0.4%) |
| Codes for special purposes (U) | 16 (1.0%) |
| External causes of morbidity and mortality (V-Y) | 40 (2.4%) |
| **Any death** | **1,640** |

N = Number of participants. ICD-10 = International Classification of Diseases, 10th revision.

### Supplemental Table 11: Underlying cause of death at five-years in the NHANES

| **Underlying cause of death (ICD-10)** | **N (%)** |
|---|---|
| Vascular death (I00-I09, I11, I13, I20-I51, and I60-I69) | 114 (30.6%) |
| Cancer death (C00-C97) | 104 (28.0%) |
| Other death | 154 (41.4%) |
| Chronic lower respiratory diseases (J40-J47) | 15 (4.0%) |
| Accidents/Unintentional injuries (V01-X59, Y85-Y86) | 12 (3.2%) |
| Alzheimer's disease (G30) | 7 (1.9%) |
| Diabetes mellitus (E10-E14) | 16 (4.3%) |
| Influenza and pneumonia (J09-J18) | 6 (1.6%) |
| Nephritis, nephrotic syndrome and nephrosis (N00-N07, N17-N19, N25-N27) | 12 (3.2%) |
| All other causes (data unavailable) | 88 (23.1%) |
| **Any death** | **372** |

N = Number of participants. NHANES = National Health and Nutrition Examination Survey. ICD-10 = International Classification of Diseases, $10^{th}$ revision.

### Supplemental Table 12: 5-year mortality risk equations

| Model | 10-year mortality risk formula |
|---|---|
| Baseline | Linear predictor = 0.3987761 x (if male) + 0.4252677 x (if 50-54) + 0.8671005 x (if 55-59) + 1.261298 x (if 60-64) + 1.61343 x (65-69) + 2.075684 x (if 70-74) + 2.407846 x (if over 75) +0.14842 x (Body mass index (per 5 kg/m2))<br>0.81978 (if smoker) + 0.07696 (SBP/10) +<br>0.13976 (if blood pressure medication) +<br>0.03922 (if cholesterol lowering medication) +<br>0.41211 (if Myocardial infarction) +<br>0.27763 (if Stroke) +<br>0.85015 (if Congestive heart failure) +<br>1.12168 (if Cancer) +<br>0.41871 (if Diabetes)<br><br>10-year mortality risk score = $0.988^{e^{\text{Linear predictor}}}$ |
| Baseline model<br>plus accelerometer data | Linear predictor =<br>0.42527 X (if male) +<br>0.41871X (if 50-54) +<br>0.85866 X (if 55-59) +<br>1.23547 X (if 60-64) +<br>1.57070 X (65-69) +<br>1.98513 X (if 70-74) +<br>2.24918 X (if over 75) +<br>0.69315 X (if smoker) +<br>0.04879 X (Body mass index (per 5 kg/m2)<br>0.07696 X (SBP/10) +<br>0.09531 X (if blood pressure medication) +<br>0.03922 X (if cholesterol lowering medication) +<br>0.37156 X (if Myocardial infarction) +<br>0.14842 X (if Stroke) +<br>0.68813 X (if Congestive heart failure) +<br>1.08181 X (if Cancer) +<br>0.32208 X (if Diabetes) +<br><br>-0.44629 X (if Daily steps/1000) +<br>0.12222 X (if Daily steps/1000) +<br>-0.08338 X (if Walking cadence per 10 steps/minute) +<br><br><br>10-year mortality risk score = $0.997^{e^{\text{Linear predictor}}}$ |
| Laboratory model | Linear predictor =<br>0.35767 X (if male) +<br>0.41211X (if 50-54) +<br>0.83725 X (if 55-59) +<br>1.24127 X (if 60-64) +<br>1.58515X(65-69) +<br>2.00013 X (if 70-74) +<br>2.28544 X (if over 75) +<br>0.63658 X (if smoker) +<br>0.12222 X (Body mass index (per 5 kg/m2)<br>0.08618 (SBP/10) +<br>0.06766 X (if blood pressure medication) +<br>0.04879 X (if cholesterol lowering medication) +<br>0.35066 X (if Myocardial infarction) + |

| | |
|---|---|
| | 0.19062 X (if Stroke) +<br>0.73716 X (if Congestive heart failure) +<br>1.06815 X (if Cancer) +<br>0.36464 X (if Diabetes) +<br><br>-0.43078 X (Albumin (per 10 g/L))<br>- 0.08338 X (ALT (per 10 U/L))<br>0.06766 X (AST (per 10 U/L) FP1)<br>- 0.02020X (AST (per 10 U/L) FP2)<br>- 0.12783 X (HDL cholesterol (per mmol/L))<br>0.13103 X (Gamma glutamyltransferase (per 100 U/L))<br>0.47000 X (Lymphocyte count (per 10^9 cells/Litre))<br>0.85015 X (Mean corpuscular haemoglobin (per 10 pg))<br>- 0.03046X (Mean corpuscular haemoglobin concentration (per 10 g/dL)<br>- 0.04082 X (Red blood count (per 10^12 cells/L) FP1)<br>0.01980 X (Red blood count (per 10^12 cells/L) FP2)<br>1.32972 X (Red blood distribution width (per 10%))<br>1.49962 X (Urea (per 100 mmol/L) FP1)<br>3.13810 X (Urea (per 100 mmol/L) FP2)<br>0.90826 X (White blood count (per 10^10 cells/L) FP1)<br>-0.26136 X (White blood count (per 10^10 cells/L) FP2)<br><br>10-year mortality risk score = $0.997^{e^{\text{Linear predictor}}}$ |
| Laboratory model plus accelerometer data | Linear predictor =<br>0.39878 X (if male) +<br>0.41211 X (if 50-54) +<br>0.82418 X (if 55-59) +<br>1.20597 X (if 60-64) +<br>1.52823 X (65-69) +<br>1.90211 X (if 70-74) +<br>2.12942 X (if over 75) +<br>0.53649 X (if smoker) +<br>0.03922 X (Body mass index (per 5 kg/m2)<br>0.08618 X (SBP/10) +<br>0.03922 X (if blood pressure medication) +<br>0.04879 X (if cholesterol lowering medication) +<br>0.32208 X (if Myocardial infarction) +<br>0.08618 X (if Stroke) +<br>0.61519 X (if Congestive heart failure) +<br>1.04028 X (if Cancer) +<br>0.29267 X (if Diabetes) +<br><br>- 0.40048X (Albumin (per 10 g/L))+<br>- 0.08338X (ALT (per 10 U/L))+<br>0.06766X (AST (per 10 U/L) FP1)+<br>- 0.02020X (AST (per 10 U/L) FP2)+<br>- 0.07257X (HDL cholesterol (per mmol/L))+<br>0.09531X (Gamma glutamyltransferase (per 100 U/L))+<br>0.38526X (Lymphocyte count (per 10^9 cells/Litre)) +<br>0.79299 X (Mean corpuscular haemoglobin (per 10 pg)) +<br>- 0.03046 X (Mean corpuscular haemoglobin concentration (per 10 g/dL)<br>- 0.04082 X (Red blood count (per 10^12 cells/L) FP1) +<br>0.01980 X (Red blood count (per 10^12 cells/L) FP2) +<br>1.19089 X (Red blood distribution width (per 10%)) + |

| | |
|---|---|
| | 1.24415 X (Urea (per 100 mmol/L) FP1)+<br>2.63261 X (Urea (per 100 mmol/L) FP2)+<br>0.74669 X (White blood count (per 10^10 cells/L) FP1) +<br>- 0.21072 X (White blood count (per 10^10 cells/L) FP2) +<br>-0.38566 X (if Daily steps/1000) +<br>0.10436 X (if Daily steps/1000) +<br>- 0.07257X (if Walking cadence per 10 steps/minute) +<br>10-year mortality risk score = $0.997^{e^{\text{Linear predictor}}}$ |

**Supplemental Table 13: Example 5-year mortality risk score of 65-year-old male , smoker, 27 BMI,160 mm Hg SBP, no blood pressure medication, Cholesterol lowering medication, prior diabetes, 7000 steps per day using the full Laboratory model plus accelerometer model**

| Risk factors | Original input value | Predictor value after required transformation | Coefficient | Product of predictor value and coefficient |
|---|---|---|---|---|
| Male | True | 1 | 0.39878 | 0.39878 |
| Age group | | | | |
| <50 years | - | 0 | 0.41211 | 0 |
| 50-54 years | - | 0 | 0.82418 | 0 |
| 55-59 years | - | 0 | 1.20597 | 0 |
| 60-64 years | - | 0 | 1.52823 | 0 |
| 65-69 years | True | 1 | 1.90211 | 1.90211 |
| 70-74 years | - | 0 | 2.12942 | 0 |
| ≥75 years | - | 0 | 0.53649 | 0 |
| Current smoker | True | 1 | 0.53649 | 0.53649 |
| Body mass index (per 5 kg/m$^2$) | 27 | 27/10 – 29.5/10 = -0.25 | 0.03922 | -0.009805 |
| Systolic blood pressure mmHg | 160 | 160/10 – 128/10 = 3.2 | 0.08618 | 0.275776 |
| Blood pressure medication | False | 0 | 0.03922 | 0 |
| Cholesterol lowering medication | True | 1 | 0.04879 | 0.04879 |
| **Prior disease** | | | | |
| Myocardial infarction | False | 0 | 0.32208 | 0 |
| Stroke | False | 0 | 0.08618 | 0 |
| Congestive heart failure | False | 0 | 0.61519 | 0 |
| Cancer | False | 0 | 1.04028 | 0 |
| Diabetes | True | 1 | 0.29267 | 0.29267 |
| **Laboratory measures** | | | | |
| Albumin (per 10 g/L) | 44.1 | 44.1/10 – 42.2/10 = 0.19000000 | -0.40048 | -0.076091200 |
| ALT (per 10 U/L) | 20.0 | 20.0/10 – 24.3/10 = -0.43000000 | - 0.08338 | 0.035853400 |
| AST (per 10 U/L) FP1 | 30 | $(30/10)^2$ - 7.541961 = 1.45803900 | 0.06766 | 0.098650919 |
| AST (per 10 U/L) FP2 | 30 | (log (30/10) X $(30/10)^2$) – 8.886379 = 1.001132 | -0.02020 | -0.020222866 |
| HDL cholesterol (per mmol/L) | 1.2 | 1.2 – 1.4 = -0.2 | -0.07257 | 0.014514000 |
| Gamma glutamyltransferase (per 100 U/L) | 32.3 | 32.3/100 – 0.3415259 = -0.01852590 | 0.09531 | -0.001765704 |
| Lymphocyte count (per 10^9 cells/Litre) | 2.6 | $2.6^{-0.5}$ - 0.7499859 = -0.1298122 | 0.38526 | -0.050011448 |
| Mean corpuscular haemoglobin (per 10 pg) | 29.2 | 29.2/10 – 33.9/10 = -0.47 | 0.79299 | -0.372705300 |
| Mean corpuscular haemoglobin concentration (per 10 g/dL) | 34.1 | $(34.1/10)^3$ –41.23892= -1.587099 | - 0.03046 | 0.048343036 |
| Red blood count (per 10^12 cells/L) FP1 | 4.8 | $4.8^3$ – 93.69396 = 16.89804 | -0.04082 | -0.689777993 |

| | | | | |
|---|---|---|---|---|
| Red blood count (per 10^12 cells/L) FP2 | 4.8 | log (4.8) X $4.8^3$ - 142.9003 = 30.57607 | 0.01980 | 0.605406186 |
| Red blood distribution width (per 10%) | 12.5 | 12.5/10 -13.5/10 = -0.1 | 1.19089 | -0.119089000 |
| Urea (per 100 mmol/L) FP1 | 5.3 | $(5.3/100)^{-0.5}$ - 4.41876 = -0.07503757 | 1.24415 | -0.093357993 |
| Urea (per 100 mmol/L) FP2 | 5.3 | log (5.3/100) – -2.957519 = 0.02005563 | 2.63261 | 0.052798652 |
| White blood count (per 10^10 cells/L) FP1 | 6.5 | $(6.5/10)^2$ - 0.4809559 = -0.0584559 | 0.74669 | -0.043648436 |
| White blood count (per 10^10 cells/L) FP2 | 6.5 | $(6.5/10)^3$ - 0.4084449 = -0.1338199 | -0.21072 | 0.028198529 |
| **Accelerometer data** | | | | |
| Daily steps (per 1000 steps) FP1 | 7000 | 7000/1000 – 9.462049 = -2.462049 | -0.38566 | 0.949513817 |
| Daily steps (per 1000 steps) FP2 | 7000 | 7000/1000 * log (7000/1000) – 22.03024 = -8.408869 | 0.10436 | -0.877549569 |
| Walking cadence (per 10 steps/minute) | 90 - 103 | 90/10 – 103/10 = -1.3 | - 0.07257 | 0.094341000 |
| $5 - year\ risk = 100 * 1 - 0.997^{e^{3.028211}}$ = 6.02 % | | | | |

### Supplemental Figure 1: Flow chart for UK Biobank participants included in analyses

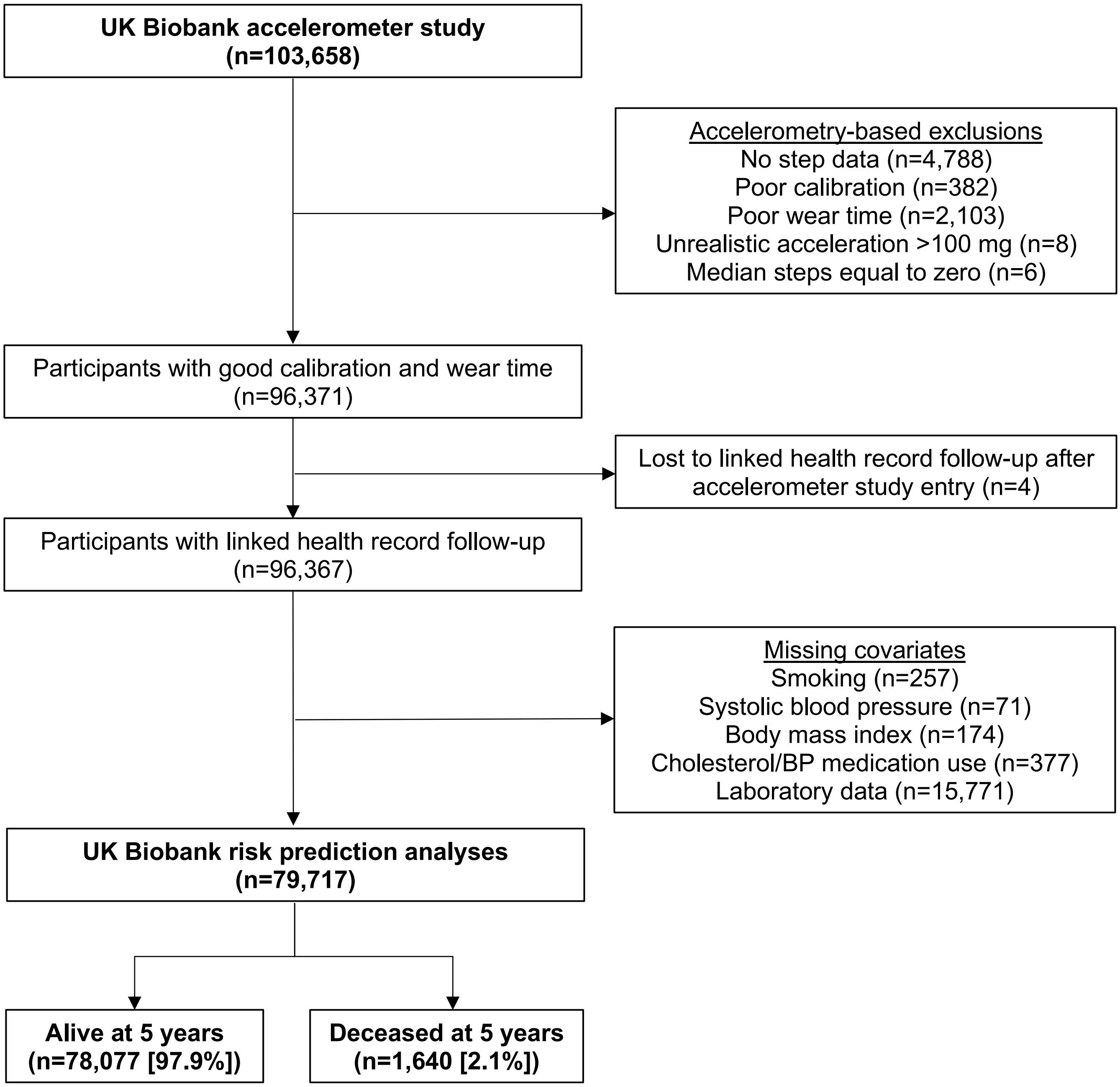


BP = Blood pressure. N = Number of participants.

**Supplemental Figure 2: Flow chart for NHANES participants included in analyses**

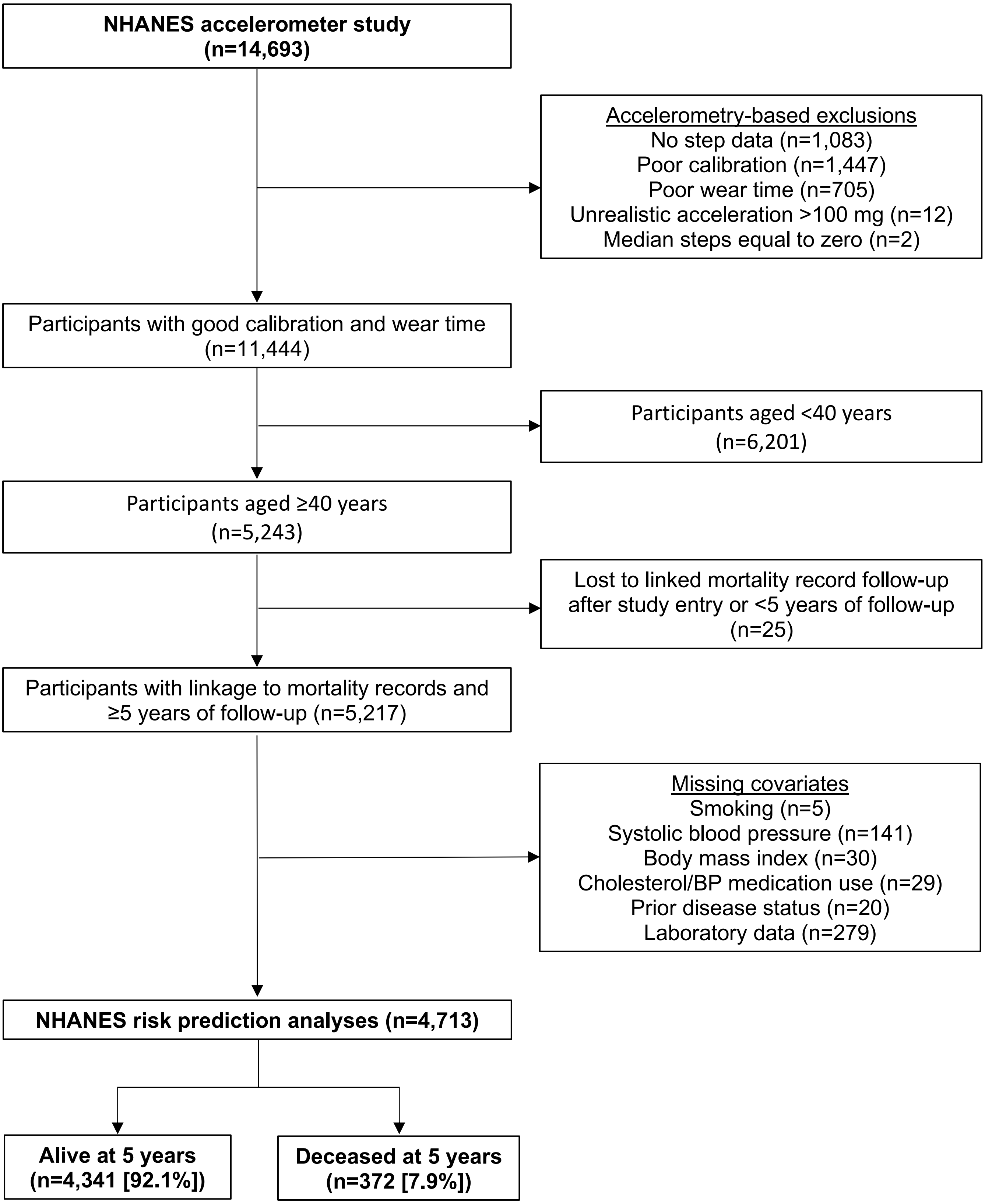


BP = Blood pressure. N = Number of participants. NHANES = National Health and Nutrition Examination Survey.

**Supplemental Figure 3: Calibration of laboratory models for 5-year all-cause mortality risk in the UK Biobank and the NHANES, by predicted risk decile**

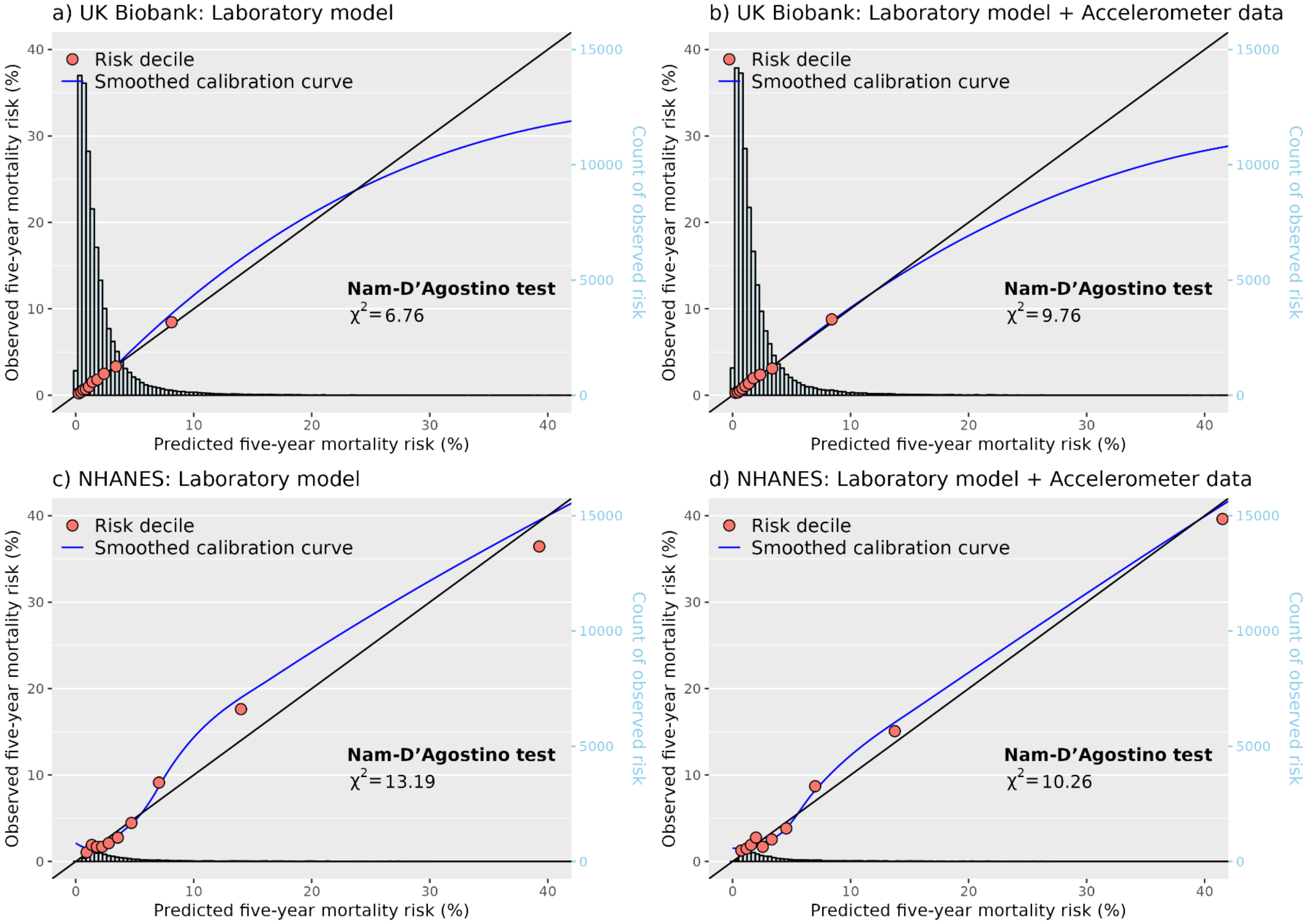


Red circles represent the observed five-year mortality risk at each decile of predicted five-year risk. Black line represents perfect calibration. Blue line represents the smoothed calibration curve of all risk deciles generated using jack-knife methods. Data for calibration plots generated using five-fold cross validation. Calibration performance estimated using Nam-D'Agostino test with nine degrees of freedom, where the smaller values represents greater performance. NHANES = National Health and Nutrition Examination Survey.

**Supplemental Figure 4: Discrimination improvement after addition of accelerometer data to baseline model for 5-year predicted mortality risk in the UK Biobank, by subgroup**

| | N | Deaths | C-index (95% CI) Baseline model | C-index (95% CI) Baseline model plus accelerometer data | Change in C-index (95% CI) |
|---|---|---|---|---|---|
| **Sex** | | | | | |
| Female | 44,410 | 640 | 0.736 (0.717, 0.756) | 0.746 (0.727, 0.765) | +0.010 (+0.004, +0.016) |
| Male | 35,307 | 1,000 | 0.758 (0.744, 0.772) | 0.768 (0.754, 0.782) | +0.010 (+0.005, +0.015) |
| **Age** | | | | | |
| <70 years | 65,021 | 960 | 0.747 (0.732, 0.763) | 0.755 (0.740, 0.770) | +0.007 (+0.003, +0.012) |
| 70+ years | 14,696 | 680 | 0.668 (0.646, 0.690) | 0.684 (0.662, 0.705) | +0.016 (+0.007, +0.026) |
| **Prior disease** | | | | | |
| Yes | 10,856 | 640 | 0.668 (0.648, 0.689) | 0.696 (0.676, 0.716) | +0.028 (+0.019, +0.039) |
| No | 68,861 | 1,000 | 0.728 (0.712, 0.743) | 0.735 (0.719, 0.750) | +0.007 (+0.002, +0.012) |
| **Overall** | **79,717** | **1,640** | **0.762 (0.752, 0.774)** | **0.770 (0.760, 0.782)** | **+0.008 (+0.005, +0.011)** |

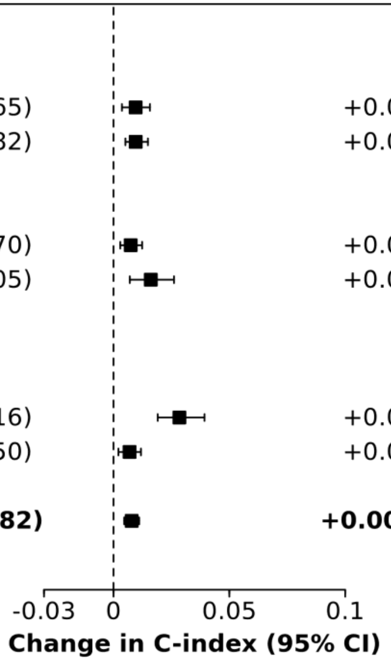


Squares represent the change in c-index compared to the baseline model and horizontal lines are 95% confidence intervals generated using 1,000 bootstrap samples. Baseline model: sex, age group, smoking, body mass index, systolic blood pressure, blood pressure medication, cholesterol medication, prior myocardial infarction, stroke, congestive heart failure, cancer, and diabetes. Accelerometer data: steps and walking cadence. Prior disease includes myocardial infarction, stroke, congestive heart failure, cancer, and diabetes. CI = Confidence interval. N = Number of participants.

**Supplemental Figure 5: Discrimination improvement after addition of accelerometer data to laboratory model for 5-year predicted mortality risk in the UK Biobank, by subgroup**

| | N | Deaths | C-index (95% CI): Laboratory model | C-index (95% CI): Laboratory and accelerometer data | Change in C-index (95% CI) |
|---|---|---|---|---|---|
| **Sex** | | | | | |
| Female | 44,410 | 640 | 0.745 (0.725, 0.764) | 0.751 (0.732, 0.770) | +0.007 (+0.002, +0.012) |
| Male | 35,307 | 1,000 | 0.772 (0.759, 0.787) | 0.778 (0.764, 0.793) | +0.005 (+0.002, +0.009) |
| **Age** | | | | | |
| <70 years | 65,021 | 960 | 0.760 (0.745, 0.776) | 0.765 (0.749, 0.781) | +0.005 (+0.001, +0.009) |
| 70+ years | 14,696 | 680 | 0.685 (0.664, 0.706) | 0.695 (0.673, 0.716) | +0.010 (+0.003, +0.017) |
| **Prior disease** | | | | | |
| Yes | 10,856 | 640 | 0.685 (0.665, 0.706) | 0.705 (0.686, 0.726) | +0.020 (+0.013, +0.029) |
| No | 68,861 | 1,000 | 0.742 (0.726, 0.756) | 0.745 (0.730, 0.760) | +0.004 (0.000, +0.008) |
| **Overall** | **79,717** | **1,640** | **0.773 (0.762, 0.785)** | **0.778 (0.767, 0.789)** | **+0.005 (+0.002, +0.007)** |

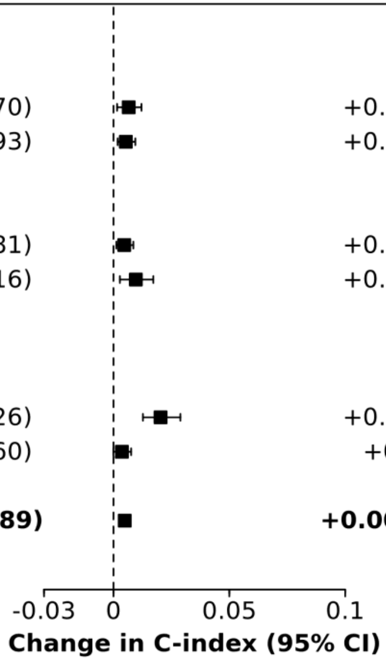


Squares represent the change in c-index compared to the laboratory model and horizontal lines are 95% confidence intervals generated using 1,000 bootstrap samples for each subgroup. Baseline model: sex, age group, smoking, body mass index, systolic blood pressure, blood pressure medication, cholesterol medication, prior myocardial infarction, stroke, congestive heart failure, cancer, and diabetes. Laboratory model: baseline model plus albumin, alanine aminotransferase, aspartate aminotransferase, high density lipoprotein cholesterol, gamma glutamyltransferase, lymphocyte count, mean corpuscular haemoglobin, mean corpuscular haemoglobin concentration, red blood cell count, red blood cell distribution width, urea, and white blood cell count. Accelerometer data: steps and walking cadence. Prior disease includes myocardial infarction, stroke, congestive heart failure, cancer, and diabetes. CI = Confidence interval. N = Number of participants.

**Supplemental Figure 6: Discrimination improvement after addition of accelerometer data to baseline model for 5-year predicted mortality risk in the NHANES, by subgroup**

| | N | Deaths | C-index (95% CI) | | Change in C-index (95% CI) |
|---|---|---|---|---|---|
| | | | Baseline model | Baseline model plus accelerometer data | |
| **Sex** | | | | | |
| Female | 2,432 | 166 | 0.801 (0.766, 0.833) | 0.820 (0.788, 0.851) | +0.019 (+0.005, +0.036) |
| Male | 2,281 | 206 | 0.797 (0.767, 0.824) | 0.808 (0.781, 0.837) | +0.012 (-0.002, +0.027) |
| **Age** | | | | | |
| <70 years | 3,593 | 130 | 0.702 (0.659, 0.747) | 0.716 (0.672, 0.762) | +0.013 (-0.012, +0.042) |
| 70+ years | 1,120 | 242 | 0.697 (0.665, 0.731) | 0.753 (0.720, 0.782) | +0.056 (+0.034, +0.080) |
| **Prior disease** | | | | | |
| Yes | 1,528 | 235 | 0.741 (0.708, 0.772) | 0.778 (0.746, 0.805) | +0.036 (+0.020, +0.053) |
| No | 3,185 | 137 | 0.778 (0.733, 0.818) | 0.785 (0.743, 0.824) | +0.007 (-0.013, +0.031) |
| **Overall** | **4,713** | **372** | **0.801 (0.778, 0.820)** | **0.816 (0.795, 0.835)** | **+0.015 (+0.007, +0.025)** |

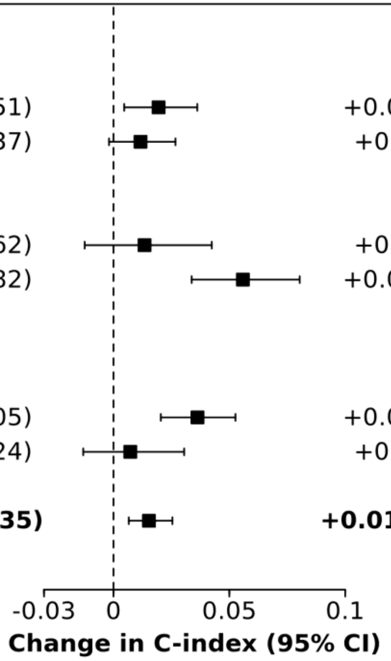


Squares represent the change in c-index compared to the baseline model and horizontal lines are 95% confidence intervals generated using 1,000 bootstrap samples. Baseline model: sex, age group, smoking, body mass index, systolic blood pressure, blood pressure medication, cholesterol medication, prior myocardial infarction, stroke, congestive heart failure, cancer, and diabetes. Accelerometer data: steps and walking cadence. Prior disease includes myocardial infarction, stroke, congestive heart failure, cancer, and diabetes. CI = Confidence interval. N = Number of participants. NHANES = National Health and Nutrition Examination Survey.

**Supplemental Figure 7: Discrimination improvement after addition of accelerometer data to laboratory model for 5-year predicted mortality risk in the NHANES, by subgroup**

| | N | Deaths | C-index (95% CI): Laboratory model | C-index (95% CI): Laboratory model plus accelerometer data | Change in C-index (95% CI) |
|---|---|---|---|---|---|
| **Sex** | | | | | |
| Female | 2,432 | 166 | 0.836 (0.808, 0.864) | 0.843 (0.815, 0.871) | +0.007 (-0.003, +0.017) |
| Male | 2,281 | 206 | 0.825 (0.799, 0.852) | 0.827 (0.801, 0.854) | +0.002 (-0.007, +0.011) |
| **Age** | | | | | |
| <70 years | 3,593 | 130 | 0.768 (0.727, 0.810) | 0.756 (0.713, 0.801) | -0.012 (-0.030, +0.006) |
| 70+ years | 1,120 | 242 | 0.736 (0.702, 0.767) | 0.770 (0.740, 0.798) | +0.034 (+0.019, +0.053) |
| **Prior disease** | | | | | |
| Yes | 1,528 | 235 | 0.786 (0.756, 0.813) | 0.803 (0.774, 0.830) | +0.018 (+0.008, +0.028) |
| No | 3,185 | 137 | 0.812 (0.775, 0.845) | 0.809 (0.774, 0.843) | -0.003 (-0.017, +0.013) |
| **Overall** | **4,713** | **372** | **0.831 (0.812, 0.850)** | **0.836 (0.816, 0.856)** | **+0.005 (-0.001, +0.011)** |

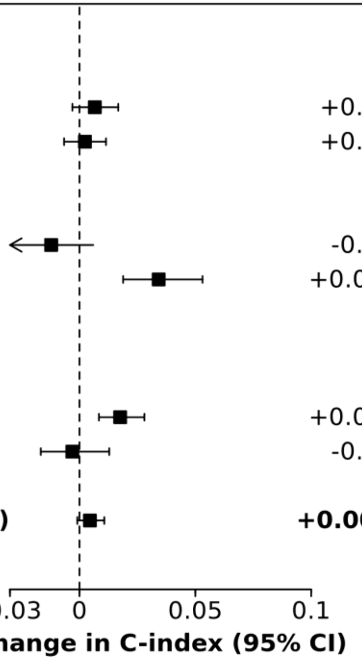


Squares represent the change in c-index compared to the laboratory model and horizontal lines are 95% confidence intervals generated using 1000 bootstrap samples for each subgroup. Baseline model: sex, age group, smoking, body mass index, systolic blood pressure, blood pressure medication, cholesterol medication, prior myocardial infarction, stroke, congestive heart failure, cancer, and diabetes. Laboratory model: baseline model plus albumin, alanine aminotransferase, aspartate aminotransferase, high density lipoprotein cholesterol, gamma glutamyltransferase, lymphocyte count, mean corpuscular haemoglobin, mean corpuscular haemoglobin concentration, red blood cell count, red blood cell distribution width, urea, and white blood cell count. Accelerometer data: steps and walking cadence. Prior disease includes myocardial infarction, stroke, congestive heart failure, cancer, and diabetes. CI = Confidence interval. N = Number of participants. National Health and Nutrition Examination Survey.

**Supplemental Figure 8: Discrimination improvement after addition of accelerometer data to baseline model for 5-year predicted mortality risk, by cause of death**

| | | C-index (95% CI) | | |
|---|---|---|---|---|
| | Deaths | Baseline model | Baseline model plus accelerometer data | Change in C-index (95% CI) |
| **UK Biobank** | | | | |
| Vascular death | 334 | 0.795 (0.773, 0.818) | 0.798 (0.776, 0.820) | +0.003 (-0.005, +0.011) |
| Cancer death | 984 | 0.747 (0.732, 0.762) | 0.751 (0.737, 0.767) | +0.004 (+0.001, +0.009) |
| Other death | 322 | 0.775 (0.750, 0.801) | 0.799 (0.773, 0.824) | +0.023 (+0.015, +0.034) |
| **All-cause mortality** | **1,640** | **0.762 (0.752, 0.774)** | **0.770 (0.760, 0.782)** | **+0.008 (+0.005, +0.011)** |
| **NHANES** | | | | |
| Vascular death | 114 | 0.854 (0.824, 0.886) | 0.864 (0.834, 0.898) | +0.010 (-0.005, +0.027) |
| Cancer death | 104 | 0.770 (0.725, 0.812) | 0.767 (0.716, 0.808) | -0.003 (-0.026, +0.019) |
| Other death | 154 | 0.782 (0.744, 0.818) | 0.813 (0.778, 0.847) | +0.031 (+0.015, +0.051) |
| **All-cause mortality** | **372** | **0.801 (0.778, 0.820)** | **0.816 (0.795, 0.835)** | **+0.015 (+0.007, +0.025)** |

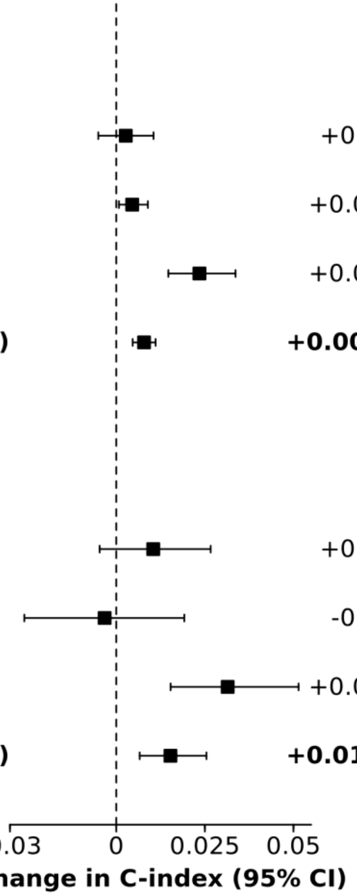


Squares represent the change in c-index comparted to the baseline model and horizontal lines are 95% confidence intervals generated using 1,000 bootstrap samples. Baseline model: sex, age group, smoking, body mass index, systolic blood pressure, blood pressure medication, cholesterol medication, prior myocardial infarction, stroke, congestive heart failure, cancer, and diabetes. Accelerometer data: daily steps, and walking cadence. See **Supplemental Tables 10 and 11** for cause of death definitions. CI = Confidence interval. N = Number of participants. NHANES = National Health and Nutrition Examination Survey.

**Supplemental Figure 9: Discrimination improvement after addition of accelerometer data to laboratory model for 5-year predicted mortality risk, by cause of death**

| | Deaths | C-index (95% CI) Laboratory model | C-index (95% CI) Laboratory model plus accelerometer data | Change in C-index (95% CI) |
|---|---|---|---|---|
| **UK Biobank** | | | | |
| Vascular death | 334 | 0.812 (0.790, 0.831) | 0.811 (0.790, 0.830) | -0.001 (-0.008, +0.006) |
| Cancer death | 984 | 0.750 (0.735, 0.766) | 0.754 (0.739, 0.769) | +0.004 (0.000, +0.008) |
| Other death | 322 | 0.802 (0.776, 0.829) | 0.816 (0.790, 0.841) | +0.014 (+0.008, +0.021) |
| **All-cause mortality** | **1,640** | **0.773 (0.762, 0.785)** | **0.778 (0.767, 0.789)** | **+0.005 (+0.002, +0.007)** |
| **NHANES** | | | | |
| Vascular death | 114 | 0.857 (0.824, 0.893) | 0.866 (0.831, 0.901) | +0.008 (-0.002, +0.020) |
| Cancer death | 104 | 0.786 (0.745, 0.825) | 0.781 (0.737, 0.820) | -0.005 (-0.020, +0.011) |
| Other death | 154 | 0.842 (0.811, 0.870) | 0.850 (0.819, 0.879) | +0.008 (-0.002, +0.019) |
| **All-cause mortality** | **372** | **0.831 (0.812, 0.850)** | **0.836 (0.816, 0.856)** | **+0.005 (-0.001, +0.011)** |

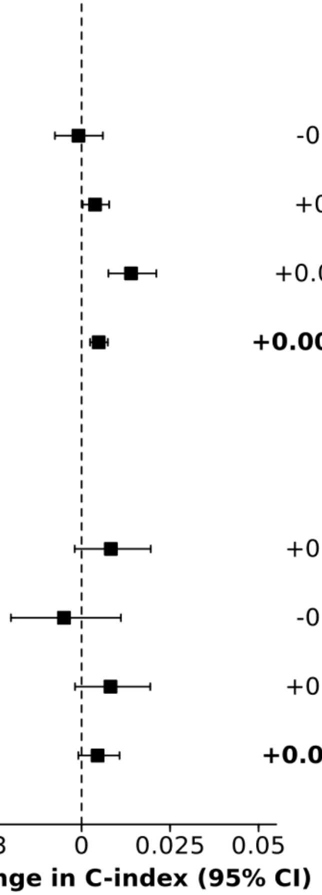


Squares represent the change in c-index comparted to the laboratory model and horizontal lines are 95% confidence intervals generated using 1,000 bootstrap samples. Baseline model: sex, age group, smoking, body mass index, systolic blood pressure, blood pressure medication, cholesterol medication, prior myocardial infarction, stroke, congestive heart failure, cancer, and diabetes. Laboratory model: baseline model plus albumin, alanine aminotransferase, aspartate aminotransferase, high density lipoprotein cholesterol, gamma glutamyltransferase, lymphocyte count, mean corpuscular haemoglobin, mean corpuscular haemoglobin concentration, red blood cell count, red blood cell distribution width, urea, and white blood cell count. Accelerometer data: daily steps, and walking cadence. See **Supplemental Tables 10 and 11** for cause of death definitions. CI = Confidence interval. N = Number of participants. NHANES = National Health and Nutrition Examination Survey.
